# Metabolism-Disrupting Chemical Mixtures during Pregnancy, Folic Acid Supplementation, and Liver Injury in Mother-Child Pairs

**DOI:** 10.1101/2024.06.13.24308903

**Authors:** Sandra India-Aldana, Vishal Midya, Larissa Betanzos-Robledo, Meizhen Yao, Cecilia Alcalá, Syam S. Andra, Manish Arora, Antonia M. Calafat, Jaime Chu, Andrea Deierlein, Guadalupe Estrada-Gutierrez, Ravikumar Jagani, Allan C. Just, Itai Kloog, Julio Landero, Youssef Oulhote, Ryan W. Walker, Shirisha Yelamanchili, Andrea A. Baccarelli, Robert O. Wright, Martha María Téllez Rojo, Elena Colicino, Alejandra Cantoral, Damaskini Valvi

**Affiliations:** Department of Environmental Medicine and Climate Science, Icahn School of Medicine at Mount Sinai, New York, NY, USA; Center for Nutrition and Health Research, National Institute of Public Health, Cuernavaca, Morelos, Mexico; Division of Laboratory Sciences, National Center for Environmental Health, Centers for Disease Control and Prevention (CDC), Atlanta, GA, USA; Department of Pediatrics, Icahn School of Medicine at Mount Sinai, New York, NY, USA; New York University School of Global Public Health, New York, NY, USA; National Institute of Perinatology, Mexico City, Mexico; Department of Epidemiology, School of Public Health, Brown University, Providence, RI, USA; Harvard T.H. Chan School of Public Health, Boston, MA, USA; Health Department, Iberoamericana University, Mexico City, Mexico

**Keywords:** Metabolism-disrupting chemicals, steatotic liver disease, folic acid, cobalt, mother-child health

## Abstract

**Background and Aims:** Scarce knowledge about the impact of metabolism-disrupting chemicals (MDCs) on liver injury limits opportunities for intervention. We evaluated pregnancy MDC-mixture associations with liver injury and effect modification by folic acid (FA) supplementation in mother-child pairs.

**Methods:** We studied ∼200 mother-child pairs from the Mexican PROGRESS cohort, with measured 43 MDCs during pregnancy (estimated air pollutants, blood/urine metals or metalloids, urine high- and low-molecular-weight phthalate [HMWPs, LMWPs] and organophosphate-pesticide [OP] metabolites), and serum liver enzymes (ALT, AST) at ∼9 years post-parturition. We defined liver injury as elevated liver enzymes in children, and using established clinical scores for steatosis and fibrosis in mothers (i.e., AST:ALT, FLI, HSI, FIB-4). Bayesian Weighted Quantile Sum regression assessed MDC-mixture associations with liver injury outcomes. We further examined chemical-chemical interactions and effect modification by self-reported FA supplementation.

**Results:** In children, many MDC-mixtures were associated with liver injury outcomes. Per quartile HMWP-mixture increase, ALT increased by 10.1% (95%CI: 1.67%, 19.4%) and AST by 5.27% (95% CI: 0.80%, 10.1%). LMWP-mixtures and air pollutant-mixtures were associated with higher AST and ALT, respectively. Air pollutant and non-essential metal/element associations with liver enzymes were attenuated by maternal cobalt blood concentrations (*p*-interactions<0.05). In mothers, only the LMWP-mixture was associated with liver injury [OR=1.53 (95%CI: 1.01, 2.28) for HSI>36, and OR=1.62 (95%CI: 1.05, 2.49) for AST:ALT<1]. In mothers and children, most associations were attenuated (null) at FA supplementation≥600mcg/day (*p*-interactions<0.05).

**Conclusions:** Pregnancy MDC exposures may increase liver injury risk, particularly in children. These associations may be attenuated by higher FA supplementation and maternal cobalt levels.

## Introduction

Steatotic liver diseases are on the rise worldwide. Over a third of the general adult population is affected by metabolic dysfunction-associated steatotic liver disease (MASLD), formerly known as non-alcoholic fatty liver disease.[1] MASLD is characterized by hepatic steatosis in conjunction with one cardiometabolic risk factor, and can progress to liver fibrosis and metabolic-dysfunction-associated steatohepatitis (MASH), increasing mortality risk.[1, 2] Latin-Americans and Hispanics are disproportionally affected and are more likely to develop MASLD and advanced fibrosis compared to non-Hispanic Whites.[3, 4] In Mexico, it is estimated that ∼20% of young adults,[5] and up to 60% of children with obesity, may have MASLD.[6]

Beyond high-fructose diets and cardiometabolic disease, higher exposure to metabolism-disrupting chemicals (MDCs) (e.g., phthalates, heavy metals, pesticides, and air pollutants) may also cause liver injury, as supported by recent epidemiological[7–10] and toxicological studies.[11, 12] Exposure to MDCs in the sensitive pregnancy period is particularly concerning for both mothers and offspring, as it may alter endocrine and metabolic systems and fetal epigenetic programming leading to long-term cardiometabolic effects and MASLD in later life. However, there is scarce knowledge about the impact of pregnancy MDC-mixture exposures on long-term liver health in mothers and children. Furthermore, emerging evidence from experimental[13] and epidemiological studies[14] suggests that folic acid (FA) supplementation may prevent or treat liver injury. FA supplementation has been shown to attenuate MDC-associations (i.e. air pollutants, phthalates) with adverse birth[15] and neurodevelopmental outcomes,[16] but its potential modifying role in MDC associations with liver injury has not been studied.

Therefore, we applied a state-of-the-art data science framework to determine the mixture effects of 43 MDCs during pregnancy on liver injury in mothers and offspring a decade later. We hypothesized that higher MDC exposures increase the risk for liver injury, and that higher FA supplementation attenuates these associations.

## Methods

### Design and Population

We used data from the prospective, population-based, Programming Research in Obesity, Growth, Environment and Social Stressors (PROGRESS) cohort which enrolled 948 mother-newborn pairs from the Mexico City area (Mexico) followed to date (enrollment criteria provided in **Supplementary Methods 1**).[17] MDC exposures were measured during pregnancy (2007-2011) and liver injury outcomes a decade later (2018-2020). In this study, we included a representative subset of 234 mothers and 205 children who had measured serum liver enzymes. This study was approved by the Institutional Review Boards of Icahn School of Medicine at Mount Sinai (US) and the National Institute of Public Health (Mexico).

### Liver Injury Assessment

Alanine transaminase (ALT), aspartate aminotransferase (AST), and gamma-glutamyltransferase (GGT) were measured in fasting blood collected from mothers and children ∼9 years after childbirth (**Supplemental Methods 2**). In the mothers, we defined risk for liver injury using established clinical non-invasive score cutoffs as primary outcomes: AST:ALT ratio <1,[18–20] Hepatic Steatosis Index (HSI) >36,[21, 22] and Fatty Liver Index (FLI) ≥60.[23] In children, we defined liver injury using the cohort’s internal 90^th^-percentile for AST (≥32.5 U/L) and ALT (≥25.3 U/L), which was comparable to the 95% percentile of NHANES children,[24] and the 25 U/L ALT-cutoff commonly used for clinical risk stratification in North American young children.[25] Primary study outcomes (ALT and AST) were also analyzed continuously to enhance statistical power and comparability between mothers and children. Scores for fibrosis in mothers (e.g., Fibrosis-4) and children (PNFS) were also calculated and considered for analysis as secondary outcomes (**Supplementary Methods 2**).

### MDC Exposures Assessment

MDC estimates (outdoor air pollutants) and biomarkers (metals/metalloids, pesticides, phthalates) in pregnant women were measured as previously detailed.[26–29]

Briefly, we estimated daily concentrations in the Mexico City Metropolitan Area at 1-km^2^ grids for two air pollutants (PM_2.5_, NO_2_) based on household location using validated spatiotemporal models and satellite information.[28, 30] We then averaged concentrations for the three pregnancy trimesters.

MDC biomarker analysis is detailed in **Supplementary Methods 3-5**. We measured 15 metals and metalloids in mothers’ urine (n=234) and 6 metals or trace elements in mothers’ blood (n=232) collected during the 2^nd^ and 3^rd^ pregnancy trimesters in trace element-free tubes.

Essential metals or trace elements may also have protective health effects, thus we evaluated the mixture associations of essential metals/trace elements vs non-essential metals/metalloids separately in analyses.[31] A subset of mothers (n=117) had 7 organophosphate pesticide (OP) metabolites measured in a single spot urine sample collected during the 2^nd^ trimester of pregnancy. Last, 15 urine phthalate metabolites were measured during the 2^nd^ and 3^rd^ pregnancy trimesters in phthalate-free tubes. For metals and phthalates, we averaged the 2^nd^ and 3^rd^ pregnancy concentrations as a proxy of exposure throughout pregnancy. MDCs with concentrations above the detection limit (LOD) for at least 60% of participants were included in statistical analyses, after substituting below LOD values by LOD/√2. Phthalates, OPs, and metals/metalloids concentrations measured in urine were corrected using specific gravity to account for urine dilution.[32]

A total of 43 MDCs were included for analyses, including 2 air pollutants, 5 OP metabolites, 7 essential metals/trace elements,14 non-essential metals or elements, 5 low-molecular-weight phthalates (LMWPs) and 10 high-molecular-weight phthalates (HMWPs). A complete list with chemical nomenclatures, biological matrices, and units is provided in **Table S3**.

### Additional Variables

Socio-demographic and lifestyle variables in PROGRESS mothers and children were measured as previously detailed.[17, 33] Our analysis accounted for household socio-economic status (SES) during pregnancy, maternal age at partum, parity, self-reported maternal passive/active smoking status and alcohol intake during pregnancy, maternal pre-pregnancy body mass index (BMI), self-reported FA intake during pregnancy (average of 2^nd^ and 3^rd^ trimesters), child’s sex, daily sedentary time in hours, sugar-sweetened beverages, and child’s exact age and puberty status (Tanner stage) at the 9-year examination. Additionally, fasting blood HbA1c,[34] whole blood platelets, plasma triglycerides (**Supplementary Methods 6**) [35] and anthropometry [waist circumference (WC) and BMI] were measured in mothers at the 9-year examination and were used to construct liver steatosis and fibrosis indexes.

### Statistical Analyses

Statistical analyses included 205 children and 234 mothers. Few outliers in liver enzyme levels were excluded using Rosner’s test[36]: n=2 for ALT, n=1 for AST, n=3 for GGT in children and n=4 for ALT or AST, and n=1 for GGT in mothers. Logarithmic transformations normalized MDC exposure (log_2_) and liver enzyme (ln) distributions. We used Bayesian Weighted Quantile Sum (BWQS) regression models to evaluate associations between MDC-mixtures during pregnancy and liver injury outcomes in mothers or children a decade later. We conducted analyses separately by chemical class and for the overall mixture containing all MDCs. All coefficients are expressed per quartile increase in the MDC-mixture. To facilitate interpretation for continuous outcomes, we re-transformed estimates from the log (base=e) scale to % change in the outcome by quartile MDC-mixture increase.

We used a novel machine-learning framework[37] that combines repeated hold-out Signed Iterative Random Forest (rh-SiRF) and regression approaches to explore potential chemical-chemical interactions within the overall MDC-mixture in association with liver injury outcomes (**Supplementary Methods 7**). Rh-SiRF analyses were conducted only for primary liver outcomes associated with the overall MDC-mixture to reduce false positive results. To validate our results, we examined whether identified interactions in one study population (either in mothers or children) were replicated in their counterpart population.

We conducted stratified analyses by FA supplementation levels of 600 µg/day, which is equivalent to the recommended folate intake during pregnancy (dietary folate equivalents), and the average recommended FA supplementation for pregnant women according to clinical guidelines (400-800 µg/day).[38–40] Almost all study participants (∼90%) reported FA supplementation above 400 µg/day. We also tested effect modification by FA supplementation by including a cross-product term between the dichotomized variable (FA≥600 µg vs. FA<600 µg/day) and the BWQS weighted index mean in unstratified models.

Covariates adjusted in statistical models were selected *a priori*, based on clinical relevance, or based on statistical significance in our dataset. Maternal liver injury models were controlled for continuous pre-pregnancy BMI, continuous age at partum, SES, parity, passive or active smoking status during pregnancy, and alcohol intake during pregnancy. Child liver injury models were adjusted for the above-mentioned covariates in addition to child’s sex and age. Childhood obesity, puberty status, sugar-sweetened beverages, and sedentary time were also evaluated in secondary analyses to confirm results. Handling of missing values and additional secondary analyses are detailed in **Supplementary Methods 8**. All analyses were conducted in Rv4.3.0. The significance level was set at a *p*-value<0.5.

## Results

### Study population description

Mothers had a mean (SD) age of 28.1 (5.4) years at partum. The majority had low SES (53%) and overweight/obesity at pre-pregnancy (57%) and at follow-up (82%) (**Table 1**). One-fifth of women reported FA supplementation above 600 µg/day. Children (52% males) had a mean (SD) age of 9.36 (0.86) years and most were at puberty (79%) (**Table 1**; pregnancy variable distributions in the subset of 205 children are provided in **Table S4**). Spearman correlations for liver enzymes between mothers and children were *ρ*=0.18 (p=0.011) for ALT and *ρ*=0.23 (p=0.001) for AST (**Table S5**). Only ∼5% of mothers had ALT and AST levels above the upper limit of normal (**Table 1**). Based on FLI and HSI scores and AST:ALT ratio, 43%, 54%, and 19% of women, respectively, were at risk for having steatosis. Based on FIB-4, APRI and NAFLD-FS scores, only 5 women were at risk for having moderate or advanced fibrosis, thus we only analyzed continuous FIB-4 in additional analyses. Stronger correlations were observed between MDCs of the same class (**Figure S1**). MDC distributions are provided in **Table S3.**

**Table 1.**
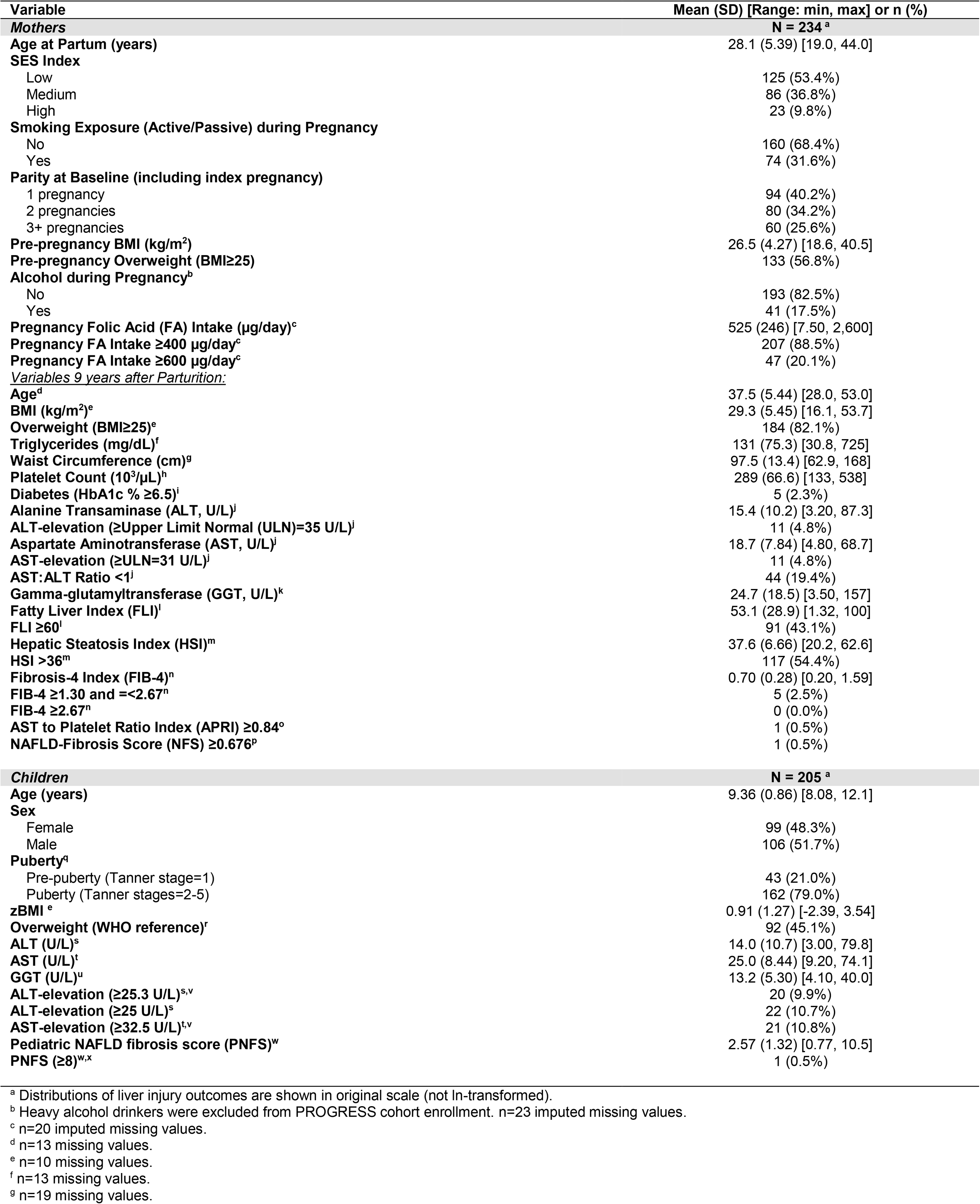
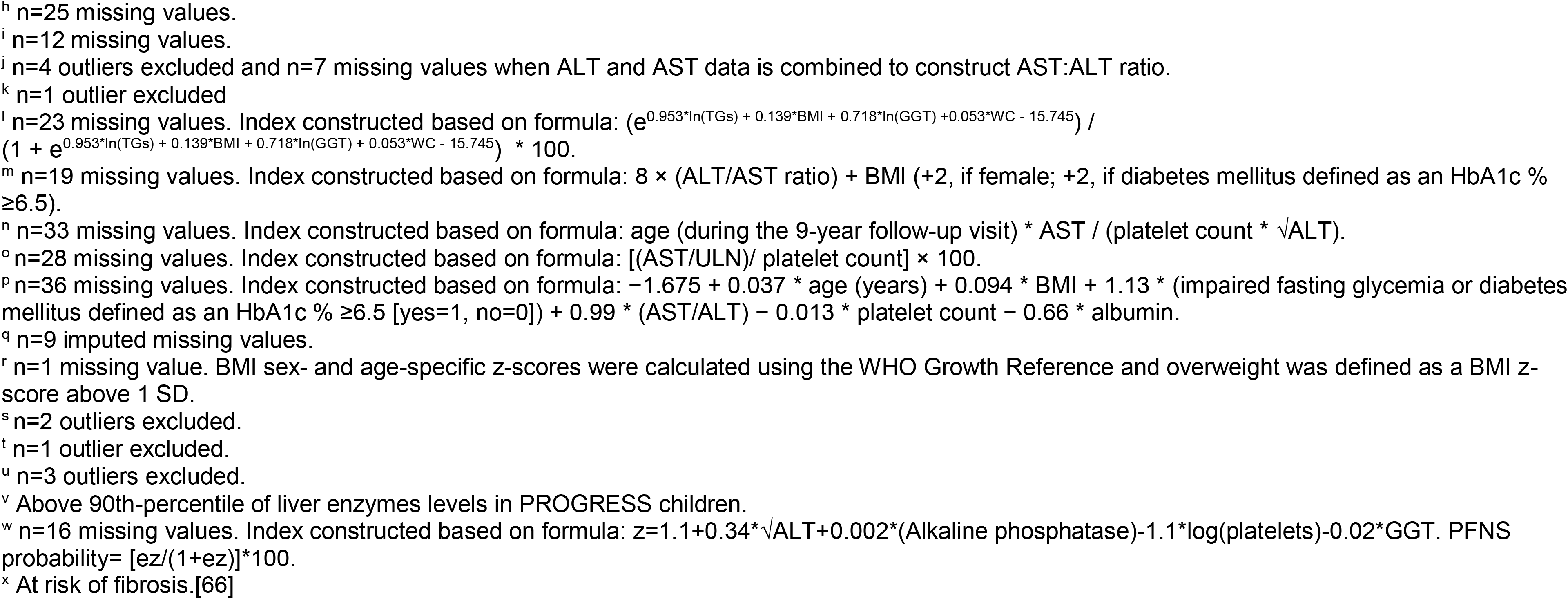
Characteristics and Outcomes in PROGRESS Mothers and Children.

| Variable | Mean (SD) [Range: min, max] or n (%) |
| --- | --- |
| **Mothers** |  |
| **N = 234<sup>a</sup>** |  |
| Age at Partum (years) | 28.1 (5.39) [19.0, 44.0] |
| SES Index |  |
| Low | 125 (53.4%) |
| Medium | 86 (36.8%) |
| High | 23 (9.8%) |
| Smoking Exposure (Active/Passive) during Pregnancy |  |
| No | 160 (68.4%) |
| Yes | 74 (31.6%) |
| Parity at Baseline (including index pregnancy) |  |
| 1 pregnancy | 94 (40.2%) |
| 2 pregnancies | 80 (34.2%) |
| 3+ pregnancies | 60 (25.6%) |
| Pre-pregnancy BMI (kg/m<sup>2</sup>) | 26.5 (4.27) [18.6, 40.5] |
| Pre-pregnancy Overweight (BMI≥25) | 133 (56.8%) |
| Alcohol during Pregnancy<sup>b</sup> |  |
| No | 193 (82.5%) |
| Yes | 41 (17.5%) |
| Pregnancy Folic Acid (FA) Intake (µg/day)<sup>c</sup> | 525 (246) [7.50, 2,600] |
| Pregnancy FA Intake ≥400 µg/day<sup>c</sup> | 207 (88.5%) |
| Pregnancy FA Intake ≥600 µg/day<sup>c</sup> | 47 (20.1%) |
| <u>Variables 9 years after Parturition:</u> |  |
| Age<sup>d</sup> | 37.5 (5.44) [28.0, 53.0] |
| BMI (kg/m<sup>2</sup>)<sup>e</sup> | 29.3 (5.45) [16.1, 53.7] |
| Overweight (BMI≥25)<sup>e</sup> | 184 (82.1%) |
| Triglycerides (mg/dL)<sup>f</sup> | 131 (75.3) [30.8, 725] |
| Waist Circumference (cm)<sup>g</sup> | 97.5 (13.4) [62.9, 168] |
| Platelet Count (10<sup>3</sup>/µL)<sup>h</sup> | 289 (66.6) [133, 538] |
| Diabetes (HbA1c % ≥6.5)<sup>i</sup> | 5 (2.3%) |
| Alanine Transaminase (ALT, U/L)<sup>j</sup> | 15.4 (10.2) [3.20, 87.3] |
| ALT-elevation (≥Upper Limit Normal (ULN)=35 U/L)<sup>j</sup> | 11 (4.8%) |
| Aspartate Aminotransferase (AST, U/L)<sup>j</sup> | 18.7 (7.84) [4.80, 68.7] |
| AST-elevation (≥ULN=31 U/L)<sup>j</sup> | 11 (4.8%) |
| AST:ALT Ratio <1<sup>j</sup> | 44 (19.4%) |
| Gamma-glutamyltransferase (GGT, U/L)<sup>k</sup> | 24.7 (18.5) [3.50, 157] |
| Fatty Liver Index (FLI)<sup>l</sup> | 53.1 (28.9) [1.32, 100] |
| FLI ≥60<sup>l</sup> | 91 (43.1%) |
| Hepatic Steatosis Index (HSI)<sup>m</sup> | 37.6 (6.66) [20.2, 62.6] |
| HSI >36<sup>m</sup> | 117 (54.4%) |
| Fibrosis-4 Index (FIB-4)<sup>n</sup> | 0.70 (0.28) [0.20, 1.59] |
| FIB-4 ≥1.30 and <2.67<sup>n</sup> | 5 (2.5%) |
| FIB-4 ≥2.67<sup>n</sup> | 0 (0.0%) |
| AST to Platelet Ratio Index (APRI) ≥0.84<sup>o</sup> | 1 (0.5%) |
| NAFLD-Fibrosis Score (NFS) ≥0.676<sup>p</sup> | 1 (0.5%) |
| **Children** |  |
| **N = 205<sup>a</sup>** |  |
| Age (years) | 9.36 (0.86) [8.08, 12.1] |
| Sex |  |
| Female | 99 (48.3%) |
| Male | 106 (51.7%) |
| Puberty<sup>q</sup> |  |
| Pre-puberty (Tanner stage=1) | 43 (21.0%) |
| Puberty (Tanner stages=2-5) | 162 (79.0%) |
| zBMI<sup>e</sup> | 0.91 (1.27) [-2.39, 3.54] |
| Overweight (WHO reference)<sup>r</sup> | 92 (45.1%) |
| ALT (U/L)<sup>s</sup> | 14.0 (10.7) [3.00, 79.8] |
| AST (U/L)<sup>t</sup> | 25.0 (8.44) [9.20, 74.1] |
| GGT (U/L)<sup>u</sup> | 13.2 (5.30) [4.10, 40.0] |
| ALT-elevation (≥25.3 U/L)<sup>s,v</sup> | 20 (9.9%) |
| ALT-elevation (≥25 U/L)<sup>s</sup> | 22 (10.7%) |
| AST-elevation (≥32.5 U/L)<sup>t,v</sup> | 21 (10.8%) |
| Pediatric NAFLD fibrosis score (PNFS)<sup>w</sup> | 2.57 (1.32) [0.77, 10.5] |
| PNFS ≥8<sup>w,x</sup> | 1 (0.5%) |

<sup>a</sup> Distributions of liver injury outcomes are shown in original scale (not ln-transformed).
<sup>b</sup> Heavy alcohol drinkers were excluded from PROGRESS cohort enrollment. n=23 imputed missing values.
<sup>c</sup> n=20 imputed missing values.
<sup>d</sup> n=13 missing values.
<sup>e</sup> n=10 missing values.
<sup>f</sup> n=13 missing values.
<sup>g</sup> n=19 missing values.

### Main associations between pregnancy MDC-mixtures and liver injury in mothers and children

In children, higher exposure to mixtures of LMWPs, HMWPs, air pollutants, and the overall MDC-mixture, were associated with increases in ALT and/or AST levels (**Figure 1**; **Figure S2**). Specifically, per 1-quartile increase in gestational HMWP biomarker-mixtures, we observed increases of 10.1% in ALT (95% CI: 1.67%, 19.4%) (**Figure 1A**) and 5.27% in AST (95% CI: 0.80%, 10.1%) (**Figure 1E**). Similarly, per quartile increase in the HMWP mixture, children had a 94% greater likelihood of having elevated ALT (OR_HMWP_ =1.94; 95% CI: 1.11, 3.56) (**Figure 1C**). Per 1-quartile increase in gestational LMWP biomarker-mixtures, AST levels increased on average by 4.98% (95% CI: 0.73%, 9.75%) (**Figure 1E**) and a marginally significant association in the same direction was also observed between LMWPs and ALT in children: 6.45% (95% CI: -1.67%, 15.5%) (**Figure 1A**). Top chemical contributors to these phthalate-mixture group associations were MECPP (10.9%) and MCOP (11.4%) for ALT (**Figures 1B, 1D; Table S6**), and MiBP (23.1%) and MCOP (11.1%) for AST (**Figure 1F**; **Table S7**). In addition, per 1-quartile increase in gestational exposure to the air pollutant-mixture, ALT levels increased by 9.66% (95% CI: 1.05%, 19.6%) in children (**Figure 1A**). Associations for non-essential metals/trace element and OP pesticide biomarker-mixtures with liver injury outcomes tended to be positive in children, but did not reach statistical significance. The overall gestational MDC exposure-mixture (all studied MDCs) was also positively associated with higher levels of ALT [β=14.6% (95% CI: 0.94%, 31.3%)] and AST [β=7.54% (95% CI: 0.52%, 15.4%)] in children (**Figure S2A**). Top chemicals contributing to the overall MDC-mixture associations with ALT and/or AST levels were PM_2.5_ (2.6%-2.9%), Cr (2.7%-2.8%), TCP (2.5%-2.7%), and MiBP (2.5%-2.6%) (**Figure S2B-S2C**; **Table S8**).

**Figure 1.**
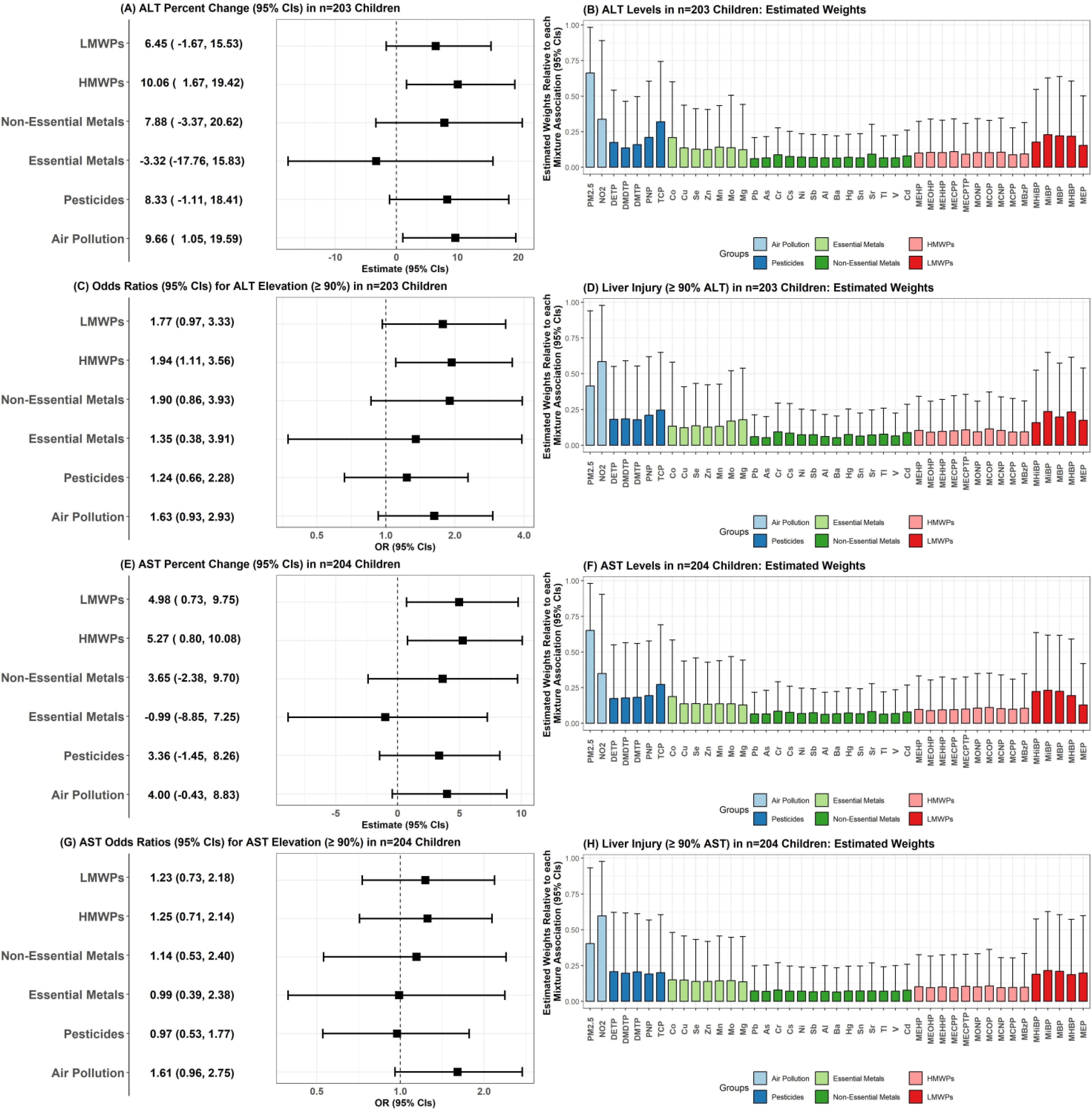
Associations Between Pregnancy MDC-Mixtures and Liver Injury Outcomes in PROGRESS Children. On the left-hand side, forest plots (A, C, E, G) represent effect estimates from BWQS, adjusted for child age, sex, and maternal continuous pre-pregnancy BMI, SES, parity, smoking exposure and alcohol intake during pregnancy, and age at parturition. Effect estimates (95% CIs) are expressed as % change in continuous outcomes or OR (95% CIs) per quartile increase in the MDC-mixtures. On the right-hand side, graphs (B, D, F, H) represent the estimated weight or contribution of each chemical to the overall MDC-mixture association.

In mothers, we found no consistent pattern of associations (**Figure 2**), with positive associations observed only for the LMWP-mixture in the overall population analysis. One quartile increase in the LMWP mixture was associated with greater likelihood of having steatosis (OR_LMWP_ =1.62; 95% CI: 1.05, 2.49 for an AST:ALT ratio below 1, and OR_LMWP_ =1.53; 95% CI: 1.01, 2.28 for an HSI>36) (**Figure 2C**). We did not find an association between pregnancy MDC-mixtures and FLI, continuous liver enzymes, or FIB-4 (**Figure S3**). There was also no association between the overall MDC-mixture and liver injury outcomes in mothers (**Figure S4**).

**Figure 2.**
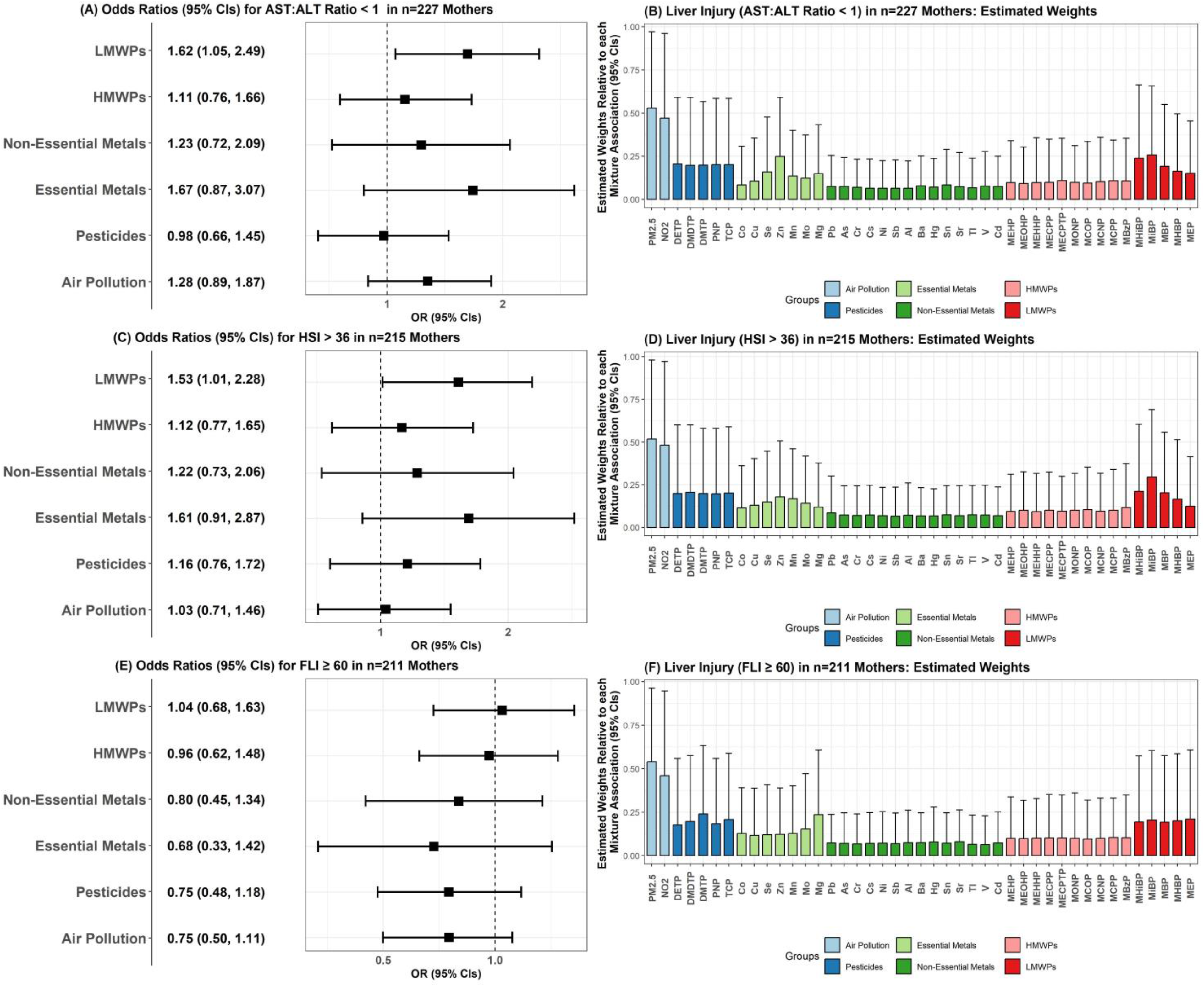
Associations Between Pregnancy MDC-Mixtures and Liver Injury Outcomes in PROGRESS Mothers. On the left-hand side, forest plots (A, C, E) represent effect estimates from BWQS, adjusted for continuous maternal pre-pregnancy BMI, SES, maternal parity, maternal smoking exposure and alcohol intake during pregnancy, and age at parturition. Effect estimates (95% CIs) are expressed as % change in OR (95% CIs) per quartile increase in the MDC-mixtures. On the right-hand side, graphs (B, D, F) represent the estimated weight or contribution of each chemical to the overall MDC-mixture association.

### Discovery analysis of potential chemical-chemical interactions associated with liver injury

We performed chemical-chemical interaction analyses in children for ALT and AST, which were associated with the overall MDC-mixture (**Figure 3**). Using the BWQS rh-SiRF algorithm, we identified 3 top two-way interactions (frequency cutoff ≥2.5%; **Supplementary Methods 5**) between Co and three other chemicals (NO_2_, Tl, MECPTP) in association with higher ALT levels in children (*p*-interaction<0.05) (**Figure 3A**). Chemical combination thresholds for two-way interactions were: lower concentrations of cobalt (Co) (≤40^th^-percentile, equal to ≤0.22 μg/L) combined with either higher levels of NO_2_ (≥35^th^-percentile, ≥29.7 μg/m^3^), or higher concentrations of Tl (≥40^th^-percentile, ≥0.34 μg/L), or lower/medium concentrations of MECPTP (≤80^th^-percentile, ≤7.1 ng/mL). We also observed 2 top two-way interactions between Co (≤80^th^ and ≤65^th^-percentiles, respectively) and either PM_2.5_ (≥70^th^-percentile) or MEP (≤50^th^-percentile) in association with higher AST levels (**Figure 3B**), as well as a two-way interaction between Cs and Sr in association with AST. A three-way interaction was also found between Co (≤80^th^-percentile), MEP (≤75^th^-percentile), and PM_2.5_ (≥70^th^-percentile) with AST levels. Moreover, we defined ‘low-low’, ‘low-high’, ‘high-low’ and ‘high-high’ exposure groups based on identified two-way chemical combinations using rh-SiRF cutoff thresholds for downstream regression analysis (**Figure 4**). These further supported that higher levels of maternal blood Co concentrations may attenuate air pollutant (NO_2_ and PM_2.5_) and Tl associations with liver injury outcomes in children, and that Cs and Sr could have synergistic effects on AST levels in children. None of the chemical-chemical interactions observed in children were evident in mothers (**Figure S5**).

**Figure 3.**
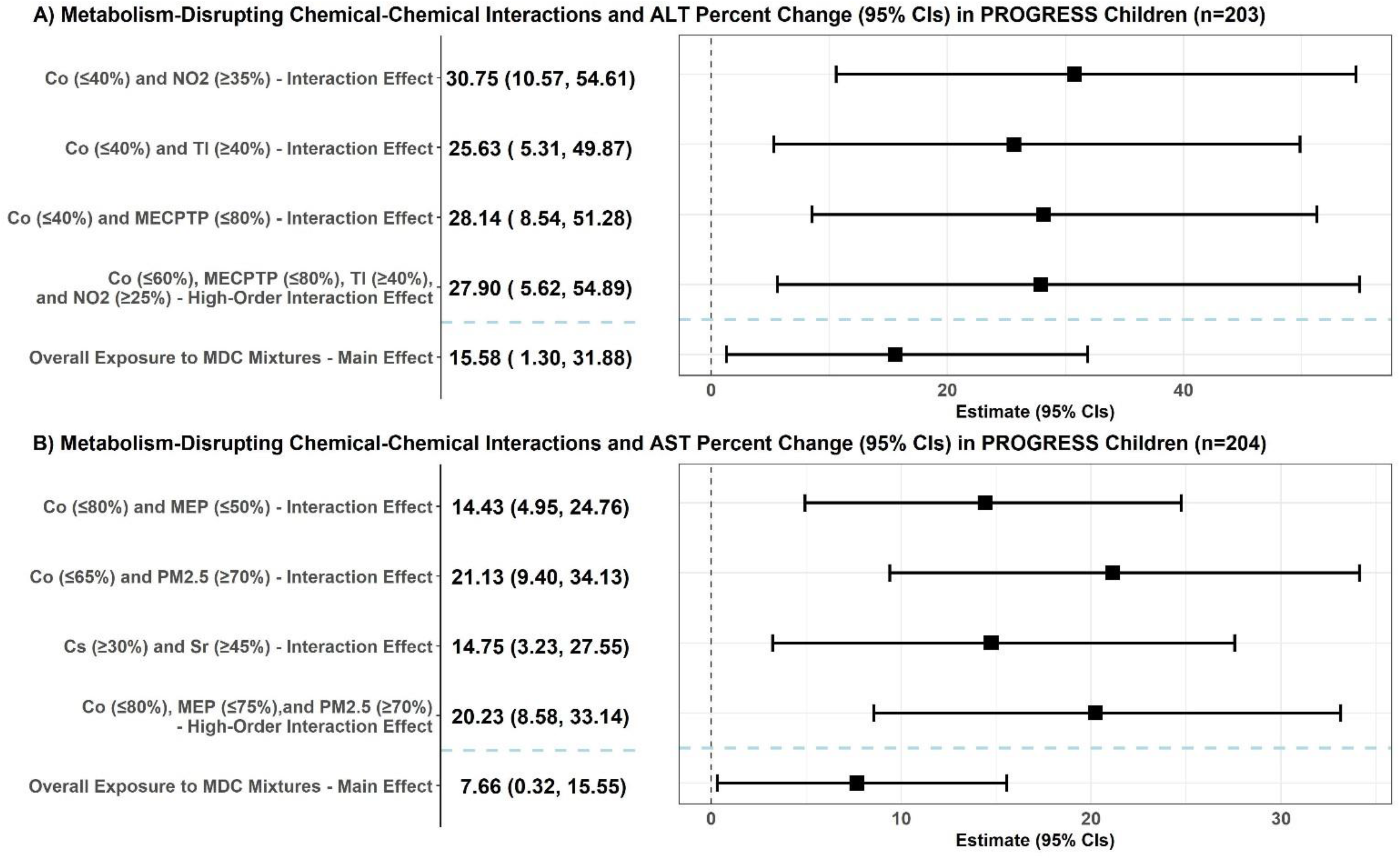
Chemical-Chemical Interactions in relation to Serum Liver Enzymes in PROGRESS Children identified using rh-SiRF. Discovered interaction indicators in association with continuous ALT and AST outcomes are shown in forest plots A and B, respectively. Effect estimates (95%CIs) are expressed as % change in the outcome per quartile increase in the interaction “cliques”. The overall MDC-mixture main effect on the outcome from BWQS without chemical-chemical interactions is also shown. Models were adjusted for child age, sex, and maternal continuous pre-pregnancy BMI, SES, parity, smoking exposure and alcohol intake during pregnancy, and age at parturition. Top chemical-chemical cliques were selected based on a frequency above 2.5% and had at least a 20% prevalence.

**Figure 4.**
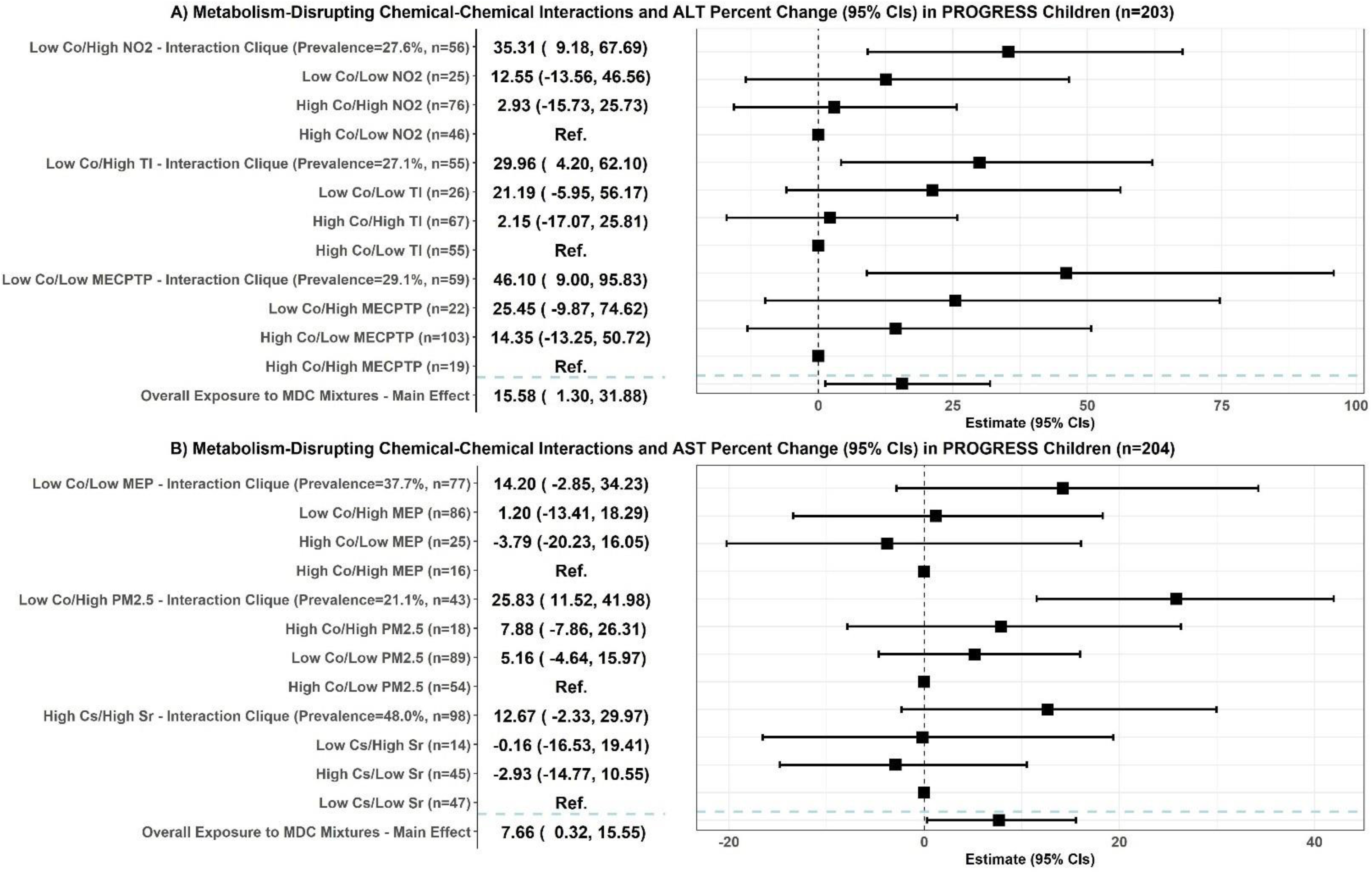
Estimates for the Associations between Two Chemical Combination Subgroups identified using rh-SiRF and Serum Liver Enzymes in PROGRESS Children. We used chemical thresholds identified by rh-SiRF (Figure 3) to define ‘low-low’, ‘low-high’, ‘high-low’ and ‘high-high’ exposure groups based on identified two-way chemical-chemical interactions. Downstream linear regression incorporated chemical combination groups as categorical predictors of continuous ALT and AST. Effect estimates (95% CIs) are expressed as % change in the outcome, adjusted for child age and sex, and maternal continuous pre-pregnancy BMI, SES, parity, maternal smoking exposure and alcohol intake during pregnancy, age at parturition, and the global BWQS MDC-mixture.

### Effect modification by FA supplementation in the associations between pregnancy MDC-mixtures and liver injury in mothers and children

In stratified analyses, we observed a consistent pattern of positive associations between MDC-mixtures and serum liver enzymes only in the subgroup of children whose mothers reported daily FA supplementation below 600 µg, but not in children whose mothers had a higher FA supplement intake (**Table 2**). Specifically, air pollutant- and OP-mixture associations with liver enzymes were positive in children at lower FA supplement intake, but tended to be negative in children with FA supplementation at or above 600 µg/day (ALT: *p*_pesticide_-interaction=0.003, *p*_air pollution_-interaction=0.043; AST: *p*_pesticide_-interaction=0.019, *p*_air pollution_-interaction=0.005). In mothers, a similar pattern of effect modification by FA supplementation was observed in the associations between air pollution and HSI (*p*_air pollution_-interaction=0.004), AST:ALT ratio (*p*_air pollution_-interaction=0.032) (**Table 3**), and continuous ALT levels (*p*_air pollution_-interaction=0.023; **Table S9**). Furthermore, higher FA supplementation during pregnancy attenuated LMWP (*p*_LMWP_-interaction=0.024) and overall MDC-mixture (*p*_Overall_-interaction=0.035) associations with the odds for having an AST:ALT ratio below 1 in mothers (**Table 3**).

**Table 2.**
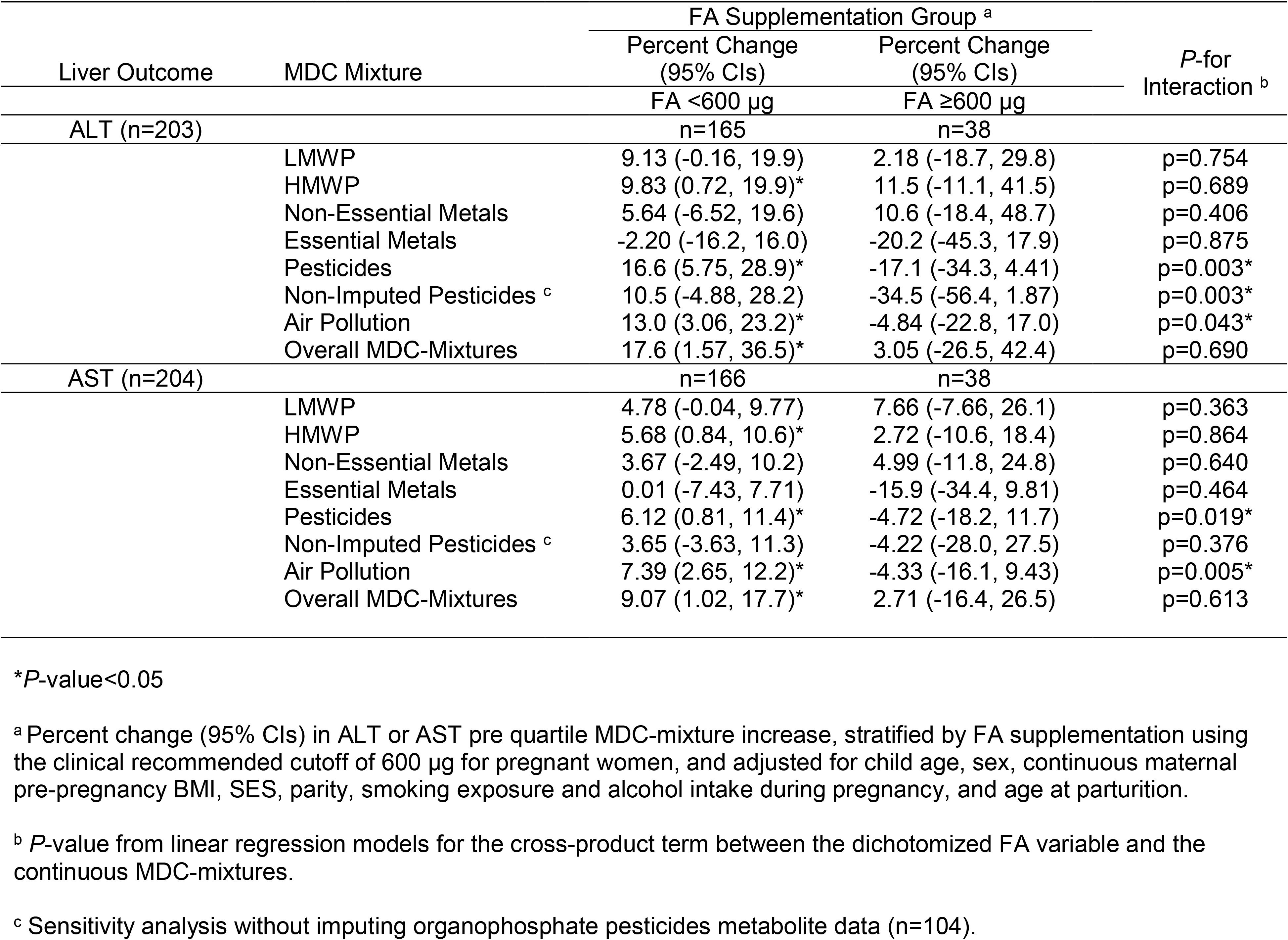
Effect Modification by Folic Acid (FA) Supplementation during Pregnancy in the Association Between MDC-Mixtures and Liver Injury in PROGRESS Children.

|  |  | FA Supplementation Group <sup>a</sup> |  | P-for<br>Interaction <sup>b</sup> |
| --- | --- | --- | --- | --- |
| Liver Outcome | MDC Mixture | Percent Change<br>(95% CIs) | Percent Change<br>(95% CIs) |  |
|  |  | FA <600 µg<br>n=165 | FA ≥600 µg<br>n=38 |  |
| ALT (n=203) |  |  |  |  |
|  | LMWP | 9.13 (-0.16, 19.9) | 2.18 (-18.7, 29.8) | p=0.754 |
|  | HMWP | 9.83 (0.72, 19.9)* | 11.5 (-11.1, 41.5) | p=0.689 |
|  | Non-Essential Metals | 5.64 (-6.52, 19.6) | 10.6 (-18.4, 48.7) | p=0.406 |
|  | Essential Metals | -2.20 (-16.2, 16.0) | -20.2 (-45.3, 17.9) | p=0.875 |
|  | Pesticides | 16.6 (5.75, 28.9)* | -17.1 (-34.3, 4.41) | p=0.003* |
|  | Non-Imputed Pesticides <sup>c</sup> | 10.5 (-4.88, 28.2) | -34.5 (-56.4, 1.87) | p=0.003* |
|  | Air Pollution | 13.0 (3.06, 23.2)* | -4.84 (-22.8, 17.0) | p=0.043* |
|  | Overall MDC-Mixtures | 17.6 (1.57, 36.5)* | 3.05 (-26.5, 42.4) | p=0.690 |
| AST (n=204) |  |  |  |  |
|  | LMWP | 4.78 (-0.04, 9.77) | 7.66 (-7.66, 26.1) | p=0.363 |
|  | HMWP | 5.68 (0.84, 10.6)* | 2.72 (-10.6, 18.4) | p=0.864 |
|  | Non-Essential Metals | 3.67 (-2.49, 10.2) | 4.99 (-11.8, 24.8) | p=0.640 |
|  | Essential Metals | 0.01 (-7.43, 7.71) | -15.9 (-34.4, 9.81) | p=0.464 |
|  | Pesticides | 6.12 (0.81, 11.4)* | -4.72 (-18.2, 11.7) | p=0.019* |
|  | Non-Imputed Pesticides <sup>c</sup> | 3.65 (-3.63, 11.3) | -4.22 (-28.0, 27.5) | p=0.376 |
|  | Air Pollution | 7.39 (2.65, 12.2)* | -4.33 (-16.1, 9.43) | p=0.005* |
|  | Overall MDC-Mixtures | 9.07 (1.02, 17.7)* | 2.71 (-16.4, 26.5) | p=0.613 |

\**P*-value<0.05
<sup>a</sup> Percent change (95% CIs) in ALT or AST pre quartile MDC-mixture increase, stratified by FA supplementation using the clinical recommended cutoff of 600 µg for pregnant women, and adjusted for child age, sex, continuous maternal pre-pregnancy BMI, SES, parity, smoking exposure and alcohol intake during pregnancy, and age at parturition.
<sup>b</sup> *P*-value from linear regression models for the cross-product term between the dichotomized FA variable and the continuous MDC-mixtures.
<sup>c</sup> Sensitivity analysis without imputing organophosphate pesticides metabolite data (n=104).

**Table 3.** Effect Modification by Folic Acid (FA) Supplementation during Pregnancy in the Association Between MDC-Mixtures with Liver Injury in PROGRESS Mothers.

| Liver Outcome | MDC Mixture | FA Supplementation Group <sup>a</sup> |  | *P*-for Interaction <sup>b</sup> |
| --- | --- | --- | --- | --- |
|  |  | OR (95% CIs) | OR (95% CIs) |  |
|  |  | FA <600 µg | FA ≥600 µg |  |
| AST:ALT <1 (n=227) |  | n=181 | n=46 |  |
|  | LMWP | 2.10 (1.30, 3.53)* | 0.82 (0.22, 2.97) | p=0.024* |
|  | HMWP | 1.41 (0.89, 2.28) | 0.48 (0.13, 1.50) | p=0.051 |
|  | Non-Essential Metals | 1.61 (0.91, 2.99) | 0.42 (0.09, 1.76) | p=0.116 |
|  | Essential Metals | 1.84 (0.84, 3.64) | 0.92 (0.11, 6.06) | p=0.346 |
|  | Pesticides | 1.11 (0.67, 1.78) | 0.48 (0.11, 1.93) | p=0.338 |
|  | Air Pollution | 1.63 (1.06, 2.66)* | 0.53 (0.19, 1.38) | p=0.032* |
|  | Overall MDC-Mixtures | 2.19 (1.10, 4.48)* | 0.32 (0.05, 1.97) | p=0.035* |
| HSI >36 (n=215) |  | n=171 | n=44 |  |
|  | LMWP | 1.47 (0.93, 2.33) | 1.72 (0.46, 6.72) | p=0.850 |
|  | HMWP | 1.13 (0.74, 1.78) | 2.37 (0.67, 9.71) | p=0.744 |
|  | Non-Essential Metals | 1.23 (0.67, 2.40) | 3.01 (0.57, 19.2) | p=0.751 |
|  | Essential Metals | 1.46 (0.70, 2.97) | 3.28 (0.60, 23.5) | p=0.370 |
|  | Pesticides | 1.15 (0.72, 1.91) | 1.28 (0.38, 4.83) | p=0.767 |
|  | Air Pollution | 1.45 (0.94, 2.34) | 0.40 (0.12, 1.07) | p=0.004* |
|  | Overall MDC-Mixtures | 1.49 (0.75, 3.09) | 4.83 (0.68, 64.8) | p=0.811 |

\**P*-value<0.05
<sup>a</sup> OR (95% CIs) for liver injury outcomes per quartile MDC-mixture increase, stratified by FA supplementation using the clinical recommended cutoff of 600 µg for pregnant women, and adjusted for continuous maternal pre-pregnancy BMI, SES, parity, smoking exposure and alcohol intake during pregnancy, and age at parturition.
<sup>b</sup> *P*-value from linear regression models for the cross-product term between the dichotomized FA variable and the continuous MDC-mixtures.

### Secondary analyses

Sex or puberty status did not modify MDC-mixture associations with liver injury outcomes in children (**Tables S10-S11**). Similarly, we found no consistent evidence for effect modification by overweight/obesity status in children (**Table S12**) or mothers (**Table S13**). Sensitivity analyses in children using an ALT cutoff of ≥25 yielded consistent results compared to primary analyses using the cohort’s 90^th^ percentile cutoff (**Table S14**). Analyses restricted to participants with available pesticide data (non-imputed dataset) yielded comparable results to the imputed pesticide analyses (**Table S15**). Lastly, adjusting for sugar-sweetened beverages and sedentary time did not meaningfully change the effect estimates for the associations between MDC-mixtures and liver injury outcomes in children (**Table S16**).

## Discussion

This is the first study to comprehensively examine the associations between MDC-mixture exposures and liver injury in mother-child pairs, and to consider potential effect modification by FA supplementation during pregnancy. Our findings showed that pregnancy exposures to air pollutants, phthalates, and/or OPs may increase the odds of liver injury, particularly for the offspring. FA supplementation above 600 µg/day during pregnancy attenuated MDC associations with liver injury in both mothers and children. Furthermore, higher maternal cobalt concentrations during pregnancy attenuated associations of air pollutants, and non-essential metal TI, with liver injury outcomes in children. These findings suggest potential benefits of nutritional interventions promoting FA and cobalamin (vitamin B_12_) intake during pregnancy (or treating folate and Co deficiencies when present) in mitigating the detrimental effects of MDCs on liver health, particularly for children. The potential palliative effects of FA and Co (or vitamin B_12_) on liver injury could be critical to inform interventions to address the current MASLD epidemic and warrant further investigation in larger clinical intervention studies.

We found associations with liver injury primarily in children compared to mothers, suggesting that the developing fetus is particularly sensitive to the hepatotoxic effects of MDCs. Few other recent studies also indicated that exposure to MDCs, particularly during gestation, may increase risk of liver injury in children and adolescents.[10, 41–43] Both our and previous studies identified associations between phthalate biomarkers and air pollutants with liver injury in children. We did not find an association between the metal/metalloid mixture and liver injury in children, contrary to previous findings in children,[10, 44] but the non-essential metals Cr and Sr were among the top contributors in our overall MDC-mixture associations. Only one previous study has examined gestational exposure to MDC-mixtures in association with pediatric liver injury.[10] This study leveraged a European pediatric population (HELIX) and found associations between phthalate mixtures and lower odds of liver injury, contrary to the positive association we found, despite comparable phthalate distributions. Differences in sociodemographic factors and potential effect modifiers (i.e. FA) may explain this discrepancy. Nevertheless, this study of predominantly White children in HELIX found positive associations with liver injury for certain metals and OPs, consistent with our findings in Mexican children.

Pregnancy MDC associations with liver injury in mothers were weaker compared to children. PROGRESS mothers were overall young with no evidence of liver fibrosis based on non-invasive screening tests. However, based on FLI and HSI scores and AST:ALT ratio, 19% to 54% of women were estimated to possibly have steatosis, a prevalence that is relatively higher than previously reported in pregnant Mexican women.[45] Only two recent cross-sectional studies have evaluated MDC-mixtures and liver injury in adults.[9, 46] One study in the US NHANES population showed a link between heavy metals, phthalates, or PFAS and MASLD. Another study in an Asian population showed higher risk of MASLD, defined by HSI and FLI, linked to a combined mixture of PFAS, phthalates, phenols, parabens, and pesticides. These findings are in agreement with our results showing an association between phthalates and elevated HSI in PROGRESS mothers.

Toxicological studies further support our findings. Metals can alter steroid receptors, increase reactive oxygen species (ROS) and oxidative stress, or induce enzymatic liver activity imbalance.[47–50] Phthalates can promote mitochondrial dysfunction, glucose metabolism disorders,[51] and lipid peroxidation.[52] OPs may induce toxicity through depletion of anti-oxidant systems.[53] Although mechanistic research involving gestational MDC exposure effects on liver injury is limited, gestational phthalate exposure has been linked to liver histological damage in rat offspring.[54] Air pollution also increases ROS and inflammation in tandem with increased liver enzymes (ALT and AST) and steatosis in *in vitro* liver cells.[55] Potential liver injury mechanisms involve upregulation of tumor-necrosis-factor-alpha which exacerbates dyslipidemia and leads to hepatic-function loss. This is consistent with our findings and previous epidemiological studies showing air-pollutant-driven deleterious liver effects.[41, 56]

The consistent novel interactions by FA and Co in the MDC associations with liver injury warrant future research, as they could have important clinical implications to mitigate MDC impact on liver health. Co is a trace essential element found in diet (e.g., green leafy vegetables, fish, meat, nuts) and an essential ring component of cobalamin (vitamin B_12_). FA (the synthetic form of vitamin B_9_/folate) is a common supplement for pregnant women important for preventing neural tube defects and poor birth outcomes. Folate and cobalamin, as methyl-nutrients, are implicated in epigenetic programming and both have been suggested as therapeutic agents to prevent or treat MASH through decreased inflammation and fibrosis in the liver of mice.[57] Similarly, Co-vitamin B_12_ deficiency increases lipid peroxidation and accumulation, branched-chain fatty acids, and histopathologic lesions, and decreases alpha-tocopherol concentrations in the liver of sheep.[58–60] In line with toxicological data, emerging epidemiological evidence suggests that patients with steatotic liver disease and/or MASLD have significantly decreased serum levels of folate and vitamin B_12_.[61–63] Overall, B vitamin supplementation has been shown to attenuate the association between ambient fine particles and epigenetic effects in epidemiological research.[64] Our findings together with prior evidence point to gestational nutritional interventions promoting FA and B_12_ vitamin supplementation, and/or treating folate and Co deficiencies when present, as potential ways to prevent long-term effects of MDCs on liver injury.

Our study had some limitations. Liver assessment was limited to non-invasive laboratory tests that have limited diagnostic accuracy. Liver imaging and/or gold-standard biopsy methods can address this limitation. Self-reported FA supplementation information is another limitation that can be addressed with integration of folate biomarker assessment in future investigations. We also did not have data on the PNPLA3 variant, which is highly frequent in Mexicans and linked to MASLD. However, given our homogenous population, we expect that >75% of our cohort has this polymorphism.[65] Lastly, the sample size of our analysis may have reduced the ability to detect associations, especially for OPs. Major study strengths included the prospective design that minimized reverse causation bias and enabled us to evaluate long-term exposure-outcome associations, the numerous MDC exposures assessed during pregnancy, the state-of-the-art data science framework we used for mixture and interaction analyses, and our study focus on a Mexican population disproportionally affected by MASLD. Additional strengths include the parallel investigation of associations in both mothers and children that allowed us to further corroborate certain findings, and the novelty of evaluating FA supplementation as a potential effect modifier in MDC-associated liver injury.

## Conclusion

In Mexican mother-child pairs, we found that pregnancy exposure mixtures to air pollutants, phthalates and OPs may increase liver injury risk later in life, particularly in children. These findings support the premise of pregnancy as a sensitive window for long-term liver health in children, but also opens an avenue of research examining pregnancy as a sensitive window for maternal liver health after parturition. Importantly, FA supplementation and higher maternal Co blood levels during pregnancy attenuated the associations between certain MDCs and liver injury, suggesting that nutritional interventions addressing folate and cobalt deficiencies in pregnant women and promoting vitamin supplementation during pregnancy could help prevent the harmful effects of MDCs in the liver. Further research is needed to fully characterize the hepatotoxic effects of MDC exposures in vulnerable populations and inform intervention strategies to address the MASLD epidemic, starting early in life, as pediatric liver injury can persist towards adulthood.

## Data Availability

All data produced in the present study are available upon reasonable request to the authors

## Author Contributions

<u>Sandra India Aldana</u>: Conceptualization, Data curation, Investigation, Formal analysis, Software, Visualization, Methodology, Writing – original draft. <u>Vishal Midya:</u> Investigation, Methodology, Writing – review & editing. <u>Larissa Betanzos-Robledo</u>: Investigation, Writing – review & editing. <u>Meizhen Yao:</u> Investigation, Methodology, Writing – review & editing. <u>Cecilia</u> <u>Alcalá</u>: Investigation, Writing – review & editing. <u>Syam Andra</u>: Investigation, Writing – review & editing. <u>Manish Arora</u>: Investigation, Writing – review & editing. <u>Antonia Calafat</u>: Investigation, Writing – review & editing. <u>Jaime Chu</u>: Investigation, Writing – review & editing. <u>Andrea</u> <u>Deierlein</u>: Investigation, Writing – review & editing. Writing – review & editing. <u>Guadalupe</u> <u>Estrada-Gutierrez</u>: Investigation, Writing – review & editing. <u>Ravikumar Jagani</u>: Investigation, Writing – review & editing. <u>Allan C. Just</u>: Investigation, Writing – review & editing. <u>Itai Kloog</u>: Investigation, Writing – review & editing. <u>Julio Landero</u>: Investigation, Writing – review & editing. <u>Youssef Oulhote</u>: Investigation, Writing – review & editing. <u>Ryan W. Walker</u>: Investigation, Writing – review & editing. Shirisha Yelamanchili: Investigation, Writing – review & editing. <u>Andrea A. Baccarelli</u>: Investigation, Writing – review & editing. <u>Robert O. Wright</u>: Funding acquisition, Investigation, Writing – review & editing. <u>Martha María Téllez Rojo</u>: Investigation, Writing – review & editing. <u>Elena Colicino:</u> Investigation, Methodology, Writing – review & editing. <u>Alejandra Cantoral Preciado</u>: Investigation, Funding acquisition, Writing – review & editing. <u>Damaskini Valvi</u>: Conceptualization, Supervision, Funding acquisition, Methodology, Writing – review & editing.

## Data Transparency Statement

Data available only upon request.

## Abbreviations

AGA: American Gastroenterological Association
Al: Aluminum
ALT: Alanine Transaminase
APRI: Aspartate aminotransferase to Platelet Ratio Index
As: Arsenic
AST: Aspartate Aminotransferase
Ba: Barium
BMI: Body Mass Index
BWQS: Bayesian Weighted Quantile Regression
Cd: Cadmium
CDC: Centers for Disease Control and Prevention
CI: Confidence Interval
Co: Cobalt
Cr: Chromium
Cs: Cesium
Cu: Copper
DEDTP: Diethyldithiophosphate
DETP: Diethylthiophosphate
DMDTP: Dimethyldithiophosphate
DMTP: Dimethylthiophosphate
FA: Folic Acid
FIB-4: Fibrosis-4 Index
FLI: Fatty Liver Index
GGT: Gamma-glutamyltransferase
HbA1c: Glycated Hemoglobin
HELIX: Human Early Life Exposome
Hg: Mercury
HMWP: High-Molecular-Weight Phthalates
HSI: Hepatic Steatosis Index
LOD: Limit of Detection
LMWP: Low-Molecular-Weight Phthalates
MASH: Metabolic Dysfunction-Associated Steatohepatitis
MASLD: Metabolic Dysfunction-Associated Steatotic Liver Disease
MBP: Mono-n-butyl phthalate
MBzP: Monobenzyl phthalate
MCNP: Mono carboxyisononyl phthalate
MCOP: Monooxononyl phthalate
MCPP: Mono-3-carboxypropyl phthalate
MDA: Malathion dicarboxylic acid
MDCs: Metabolism-Disrupting Chemicals
MECPP: Mono-2-ethyl-5-carboxypentyl phthalate
MECPTP: Mono-2-ethyl-5-carboxypentyl terephthalate
MEHHP: Mono-2-ethyl-5-hydroxyhexyl phthalate
MEHP: Mono-2-ethylhexyl phthalate
MEOHP: Mono-2-ethyl-5-oxohexyl phthalate
MEP: Monoethyl phthalate
Mg: Magnesium
MHBP: Mono-hydroxybutyl phthalate
MHiBP: Mono-hydroxyisobutyl phthalate
MiBP: Mono-isobutyl phthalate
Mn: Manganese
Mo: Molybdenum
MONP: Monooxononyl phthalate
NAFLD: Nonalcoholic Fatty Liver Disease
NAFLD-FS: NAFLD Fibrosis Score
Ni: Nickel
NO_2_: Nitrogen Dioxide
OP: Organophosphate
OR: Odds Ratio
Pb: Lead
PM_2.5_: Particulate Matter 2.5
PNFS: Pediatric NAFLD Fibrosis Score
PNP: Nitrophenol
PROGRESS: Programming Research in Obesity, Growth, Environment and Social Stressors
ROS: Reactive Oxygen Species
Sb: Antimony
SD: Standard Deviation
Se: Selenium
SES: Socio-economic Status
rh-SiRF: Repeated hold-out Signed Iterative Random Forest
Sn: Tin
Sr: Strontium
TCP: 3,5,6-Trichloro-2-Pyridinol
Tl: Thallium
ULN: Upper Limit of Normal
V: Vanadium
WC: Waist Circumference
WHO: World Health Organization
Zn: Zinc

## Acknowledgments

We thank all the PROGRESS participating families for their generous collaboration, and all the PROGRESS fieldworkers, health professionals, and researchers for their valuable dedication and contribution to this study.

This work was supported in part through the computational and data resources and staff expertise provided by Scientific Computing and Data at the Icahn School of Medicine at Mount Sinai and supported by the Clinical and Translational Science Award (CTSA) grant UL1TR004419 from the National Center for Advancing Translational Sciences.

