## Supplementary Material for "Metabolism-Disrupting Chemical Mixtures during Pregnancy, Folic Acid Supplementation, and Liver Injury in Mother-Child Pairs"

**Table of Contents**

**Supplementary Methods 1.** PROGRESS Data Framework and Inclusion and Exclusion Criteria.

**Supplementary Methods 2.** Liver Enzyme Assessment in PROGRESS and Additional Liver Injury Classifications in Mother-Child Pairs.

**Supplementary Methods 3.** Pregnancy Trace Element Measurements in Blood and Urine.

**Supplementary Methods 4.** Pregnancy Pesticide Metabolites Measured in Urine.

**Supplementary Methods 5.** Pregnancy Phthalate and Plasticizer Metabolites in Urine.

**Supplementary Methods 6.** Biomarker Laboratory Analyses in PROGRESS for Triglycerides, HbA1c, and Platelet Count.

**Supplementary Methods 7**. Chemical-chemical Interactions using repeated hold-out Signed Iterative Random Forest (rh-SiRF).

**Supplementary Methods 8.** Handling Missing Data and Additional Sensitivity Analyses Methods.

**Table S1.** Baseline Participant Characteristics in PROGRESS (n=948 Mothers) and Subset with Liver Enzyme Data (n=234 Mothers).

**Table S2.** PROGRESS Liver Enzymes Among Two Pesticide Group Subsets.

**Table S3.** Chemical Exposure Distributions for Analyses in PROGRESS.

**Table S4.** Characteristics and Outcomes in PROGRESS Children.

**Table S5.** Liver Enzyme Correlations PROGRESS.

**Figure S1.** Chemical Distributions and Correlations in PROGRESS Participants.

**Table S6.** Estimated Posterior Weights of HMWP-Mixture Group on ALT using BWQS Models in PROGRESS Children.

**Table S7.** Estimated Posterior Weights of Phthalate-Mixture Group on Continuous AST using BWQS Models in PROGRESS Children.

**Figure S2.** Global (Overall) BWQS with All Metabolism-Disrupting Chemicals in Relation to Liver Injury in PROGRESS Children.

**Table S8.** Estimated Posterior Weights of Mixture Groups on Liver Injury using Overall BWQS Models in PROGRESS Children.

**Figure S3.** Associations Between Pregnancy MDC Mixtures and Liver Injury in PROGRESS Mothers (% Change [95% CIs] per Quartile Increases in MDC Mixtures).

**Figure S4.** Global (Overall) BWQS with All Metabolism-Disrupting Chemicals in Relation to Liver Injury in PROGRESS Mothers.

**Figure S5.** Rh-SiRF Analysis for Pre-selected Two-Order Chemical-Chemical Interactions in Relation to Liver Enzymes in PROGRESS Mothers.

**Table S9.** Effect Modification by Folic Acid Supplementation during Pregnancy in the Association Between MDC Mixtures with Liver Enzymes in PROGRESS Mothers.

**Table S10.** Effect Modification by Puberty at Follow-Up Year ~9 in the Association of MDC Mixtures with Liver Injury in PROGRESS Children.

**Table S11.** Effect Modification by Sex in the Association Between MDC Mixtures with Liver Injury in PROGRESS Children.

**Table S12.** Effect Modification by Overweight/Obesity at Year ~9 in the Association Between MDC Mixtures with Liver Injury in PROGRESS Children.

**Table S13.** Effect Modification by Pre-Pregnancy Obesity Status in the Association Between MDC Mixtures with Liver Injury in PROGRESS Mothers.

**Table S14**. Sensitivity BWQS Analyses for the Associations Between Pregnancy MDC Mixtures and Liver Injury (ALT≥25) in PROGRESS Children (OR [95% CIs] per Quartile Increases in MDC Mixtures).

**Table S15.** Sensitivity BWQS Analyses for the Associations Between Pregnancy Pesticide Mixture and Liver Enzymes in PROGRESS Restricted to Participants with Measured Pesticide Data (% Change [95% CIs] per Quartile Increases in Pesticide Mixtures).

**Table S16.** Sensitivity BWQS Analyses for the Associations Between Pregnancy MDC Mixtures and Liver Injury in PROGRESS Children controlling for Lifestyle Factors (% Change or OR [95% CIs] per Quartile Increases in MDC Mixtures).

**Supplementary Methods 1. PROGRESS Data Framework and Inclusion and Exclusion Criteria.**

All mothers and their respective children were affiliated with the Mexican Social Security Institute (*Instituto Mexicano del Seguro Social* or IMSS). Women were considered for inclusion if they were pregnant, were ≥18 years of age, were at 12-20 weeks of gestation at enrollment, and were exempt from chronic conditions, including a history of infertility, heart or renal disease, diabetes, psychosis or use of anti-epilepsy drugs, use of steroids, consumption of one or more alcoholic drinks per day, or drug addiction at recruitment. Information on socio-demographic, lifestyle, and environmental factors was collected through validated questionnaires, along with biological samples during pregnancy and later examinations. Health outcomes during follow-up were measured in children and their mothers using standardized protocols. Our subset population had similar baseline characteristics as compared to the main PROGRESS cohort with the 948 mother-child pairs initially enrolled (**Table S1**). All participants enrolled in the study provided informed consent in Spanish.^1^

**Supplementary Methods 2. Liver Enzyme Assessment in PROGRESS and Additional Liver Injury Classifications in Mother-Child Pairs.**

Fasting venous blood samples were centrifuged to separate the serum at the laboratory of the National Institute of Perinatology (Mexico City). Liver enzyme biomarkers including alanine transaminase (ALT), aspartate aminotransferase (AST), and gamma-glutamyltransferase (GGT) were measured in serum samples (U/L), using a Respons®910 automated analyzer (DiaSys Diagnostic Systems, Holzheim, Germany).

In the mothers, we also calculated the proportion of mothers at risk of having advanced fibrosis based on the Fibrosis-4 Index (FIB-4),^2^ the aspartate aminotransferase to platelet ratio index (APRI),^3^ or the NAFLD fibrosis score (NAFLD-FS)^4^ using clinical cutoffs recommended by the American Gastroenterological Association (AGA). Based on non-invasive scores, less than 5% of mothers were classified as being at risk of advanced fibrosis; therefore, we only used the FIB-4 score continuously and as a secondary outcome in our analysis.

In the children, we further analyzed ALT and AST levels as continuous outcome variables to enhance statistical power and for direct comparability of results between mothers and children. The probabilities of MASLD and fibrosis in children participants were defined based on an ALT cutoff of ≥25 U/L^5^ and a pediatric NAFLD fibrosis score (PNFS) ≥8,^6^ respectively.

**Supplementary Methods 3. Pregnancy Trace Element Measurements in Blood and Urine.**

A total of 15 metals and metalloids were measured in mothers’ urine (n=234; Mg, Mo, As, Cr, Cs, Ni, Sb, Al, Ba, Hg, Sn, Sr, Tl, V, Cd) and 6 metals or trace elements were measured in mothers’ blood (n=232; Se, Zn, Co, Cu, Mn, Pb)^7^ during the 2^nd^ and 3^rd^ trimesters of pregnancy. Briefly, digested blood samples and acidified diluted urine were analyzed using external calibration using the Agilent 8800 ICP Triple Quad in MS/MS mode and QA/QC procedures were performed:

Trace element quantification of metals and metalloids was performed in the Lautenberg Environmental Health Sciences Laboratory at the Icahn school of Medicine at Mount Sinai. In brief, 200 µl of urine sample was diluted to 10 ml with diluent solution (containing 0.5 % HNO_3_, 0.005% Triton X-100 and a mixture of internal standards) in a Polypropylene trace metal free Falcon tubes (VWR® Metal-Free Centrifuge Tubes). Whole blood samples were acid digested, 1ml of blood was digested with 1 ml of concentrated HNO_3_ and 1ml of H_2_O_2_ at room temperature for 48h and brought up to 10 ml with doubly deionized water. All sample preparation work was performed in an ISO Cass 5 laminar flow clean hood in the ISO Class 6 clean room. Samples were analyzed using matrix matched calibration standards using Agilent 8900 ICP Triple Quad mass spectrometer (ICP-QQQ) (Agilent technologies, Inc., Delaware, USA) in MS/MS mode with appropriate cell gases to eliminate molecular ion interferences. Internal standards (for instance, tellurium (Te) for As and Se, Rhodium (Rh) for Cd, or lutetium (Lu) for Pb) were used to correct for the differences in sample introduction, ionization, and reaction rates in the plasma and the reaction cell. Quality control (QC) and quality assurance (QA) procedures will include analyses of initial calibration standards in the range of 0.01 ng/ml to 50 ng/ml, verification standards (ICVS) and continuous calibration verification standards (CCVS) (mixed element standards at two different concentration levels, procedural blanks, duplicates and in-house pooled urine sample at 3 levels (IHU or IHB) to monitor the accuracy, recovery rates and reproducibility of the procedure for each analytic batch. CCVS and in-House pooled samples were run after analysis of every 10 samples. All Lab recovery rates for QC by this method are 90 to 110%, inter-day and intra-day precision (given as %RSD) is <6% for samples with concentrations > LOQ. The limits of detection for Analytes by this procedure were between 0.05 ng/ml to 0.2 ng/ml^-1^ and Limit of Quantitation ranged between 0.5 ng/ml to 2 ng/ml. The laboratory analyzes National Institute of Standards and Technology’s (NIST) standard reference material (SRM) 2668 (Toxic elements in Frozen urine, Gaithersburg, MD) and NIST SRM 955c (toxic metals in Caprine blood) every three months or as needed, based on the recovery of the in-house materials. The laboratory successfully participates in the New York State, Department of Health and Center de Toxicologie-INSPQ, Quebec, Canada external performance testing programs to continuously validate this method. For statistical modeling, we computed the mean across 2^nd^ and 3^rd^ trimester specific gravity-corrected metal measurements to address variability (after correcting for dilution for samples that were in urine).

**Supplementary Methods 4. Pregnancy Organophosphate Metabolites Measured in Urine.**

Samples were analyzed at the Lautenberg Laboratory at Icahn School of Medicine at Mount Sinai with isotope-dilution liquid chromatography and tandem mass spectrometry operated in electrospray negative mode for ionization and multiple reaction monitoring for quantification. Analyses were performed in two phases. Initially, a multiclass pesticides assay implementing a single sample preparation based on the CDC method ^8,9^ with a modification was used to measure OP pesticide metabolites in a subset of 81 mothers included in the present analysis.^10^ In the second phase, using the same analytical method and instrumentation, a more extended list of pesticides (beyond the above-mentioned overlapping OPs) was quantified in an additional subset of 36 mothers, also included in our analysis. This population subset had similar liver outcome distributions compared to the subset analyzed during the first phase of the study (**Table S2**). Non-persistent pesticides and metabolites including organophosphate (OP) metabolites (DEDTP, DETP, DMDTP, DMTP, MDA, PNP, TCPY) were measured in maternal spot urine samples collected during the 2^nd^ trimester pregnancy visit. Samples were stored at -80֯C in 2mL aliquots. OP analysis were conducted in two phases using the same analytical methods and instrumentation:

We used a multiclass pesticides assay using a single sample preparation/cleanup based on the Centers for Disease Control and Prevention (CDC) method ^8,9^ with a modification;^10^ samples were analyzed using an isotope-dilution liquid chromatography and tandem mass spectrometry^8,9^ and we made a few small changes^10^ to make the method more accurate and useful for measuring a number of metabolites of organophosphorous pesticides and pyrethroid insecticides, among others, totaling 100 parent compounds and metabolites belonging to ten chemical classes of environmental contaminants exposures of public health relevance. In brief, ^13^C_12_, ^13^C_6_, ^13^C_2_, D_10_, or D_6_ labeled internal standards were added to each sample; metabolites were deconjugated with β-glucuronidase/arylsulfatase from *Helix pomatia* (product # 10127060001, Roche Diagnostics through Sigma Aldrich), followed by solid-phase extraction with an Oasis HLB hydrophilic-lipophilic balanced reversed-phase 96-well plate (30 mg sorbent per well, 30 µm particle size; Waters Corporation, Milford, MA). Low-volume sample aliquoting (0.2 mL) and cleanup procedures were automated using a liquid handler (epMotion 5075vtc; Eppendorf, Hauppauge, NY). The LC-MS/MS (ExionLC Series UHPLC and SCIEX 7500 triple quadruple MS with QTRAP enabled, AB Sciex; Framingham, MA) was operated concurrently in an electrospray positive and negative mode for ionization and multiple reaction monitoring (MRM) for quantification. Chromatographic separation was achieved on a Thermo Hypersil Gold AQ 100 x 3.0 mm analytical column with a 10 x 4.0 mm guard cartridge (Thermo Scientific, Waltham, MA). Mobile phase A was LC/MS-grade water with 0.1% acetic acid, and mobile phase B was LC/MS-grade acetonitrile and methanol (1:1 v/v). Quality controls (QC) accounted for 20% of each batch. QC included in each batch were procedural and instrumental blanks, matrix spikes in the lower, middle, and upper range of assay validation, CHEAR A and B QC urine pools, and archived proficiency testing material. Batchwise relative standard deviations (RSDs) of QCs during analysis of the study specimens were <20% for target analytes in reference or fortified material, except for analytes at or below LOQ (LOQ = 3xLOD), where RSDs up to 30% were accepted. Intra-batch precision (CV) was below 10%, and inter-batch precision was below 20% for QC analytes above the LOQ. Recoveries in batch QC were between 70% and 130%. The Mount Sinai lab’s multiclass assay for urinary pesticides and metabolites has been consistently successful in meeting acceptance criteria in the proficiency-testing programs conducted by the German External Quality Assessment Scheme (G-EQUAS) (https://app.g-equas.de/web/) ^11^ and the *Centre de Toxicologie du Québec* (CTQ)-OSEQAS: External Quality Assessment Scheme for Organic Substances in Urine (https://www.inspq.qc.ca/en/ctq/eqas/oqesas/description).^12^ The QA and QC protocols for the HHEAR/CHEAR lab assays are outlined in detail elsewhere.^13^

**Supplementary Methods 5. Pregnancy Phthalate and Plasticizer Metabolites in Urine.**

Urine phthalate metabolites were quantified using online solid phase-extraction isotope dilution high-performance liquid chromatography tandem mass spectrometry.^14^ In urine, we measured 15 phthalate metabolites^14,15^ (MHBP, MBP, MEP, MHiBP, MiBP, MECPTP, MONP, MBzP, MCNP, MCOP, MCPP, MECPP, MEHHP, MEHP, MEOHP) at two-time points, during the 2^nd^ and 3^rd^ trimesters of pregnancy. Given the relatively short elimination half-lives of these chemicals in humans (hours) we used the average of the two measurements as a better proxy of exposure during pregnancy. We classified phthalate metabolites as high-molecular-weight phthalates (HMWP) including MEHP, MEOHP, MEHHP, MECPP, MECPTP, MONP, MCOP, MCNP, MCPP, and MBzP, as well as low-molecular-weight phthalates (LMWP) including MHiBP, MiBP, MBP, MHBP, and MEP.^16^

Phthalate and 1,2-Cyclohexane dicarboxylic acid diisononyl ester (DINCH) metabolites were measured in maternal urine samples collected at the 2^nd^ and 3^rd^ pregnancy trimester visits. Urine was analyzed at the Centers for Disease Control and Prevention in December 2017 using online solid phase extraction-isotope dilution high-performance liquid chromatography tandem mass spectrometry to quantify two DINCH metabolites and 15 phthalate metabolites as described in detail previously.^14,17^ The phthalate metabolites measured were mono n-butyl phthalate (MBP), mono-isobutyl phthalate (MiBP), mono-hydroxybutyl phthalate (MHBP), mono-hydroxyisobutylphthalate (MHiBP), mono-3-carboxypropyl phthalate (MCPP), monoethyl phthalate (MEP), mono-2-ethyl-5-carboxypentyl phthalate (MECPP), mono-2-ethylhexyl phthalate (MEHP), mono-2-ethyl-5-hydroxyhexylphthalate (MEHHP), mono-2-ethyl-5-oxohexyl phthalate (MEOHP), monobenzyl phthalate (MBzP), mono(carboxy-isononyl) phthalate (MCNP) mono(carboxy-isooctyl) phthalate (MCOP), monooxononylphthalate (MONP), and mono-2-ethyl-5-carboxypentyl terephthalate (MECPTP). The DINCH metabolites measured were cyclohexane-1,2-dicarboxylic acid monohydroxy isononyl ester (MHiNCH) and cyclohexane-1,2-dicarboxylic acid mono (carboxyoctyl) ester (MCOCH). Limits of detection (LODs) ranged from 0.2 to 1.2 ng/mL, depending on the metabolite. The analysis of urine samples at the Centers for Disease Control and Prevention (CDC) laboratory did not constitute engagement in human-subjects research.

**Supplementary Methods 6. Biomarker Laboratory Analyses in PROGRESS for Triglycerides, HbA1c, and Platelet Count**.

Triglyceride levels were measured using enzymatic photometric assays on a Respons®910 automated analyzer (DiaSys Diagnostic Systems, Holzheim, Germany), glycated hemoglobin (HbA1c %) was measured on an InnovaStar analyzer (DiaSys Diagnostic Systems, Holzheim, Germany), and platelet count was measured using a Coulter AcT 5diff (Beckman Coulter, CA, USA) at the National Institute of Perinatology in Mexico.^18,19^

**Supplementary Methods 7. Chemical-chemical Interactions using repeated hold-out Signed Iterative Random Forest (rh-SiRF).**

The rh-SiRF method allows the identification of potential toxicologically-mimicking interactions on top of high-dimensional environmental chemical mixtures.^20-22^ This method uses a two-stage approach. In the first stage, we treated the residuals extracted from the BWQS mixture model as the outcome and chemical exposures as the predictors. Since the BWQS model did not account for any interactions, the residuals are assumed to contain the effect of several interactions. This algorithm identifies non-linear and multi-order combinations of exposures that are potentially predictive of the outcome. The algorithm was repeated 500 times with a training-testing split of 60%-40%. Among all possible combinations of exposures, rh-SiRF chooses the top “stable” or frequently occurring combinations among all possible combinations. In the second stage, the chosen combinations of chemical exposures were converted into non-linear interactions that identify potentially specific high-risk subgroups. A quantile-based threshold-finding algorithm was implemented to transform the exposure combinations into indicators using their threshold concentrations (expressed as specific percentiles of this cohort). The chosen combinations of chemical exposures were then converted into “chemical-cliques” that defined specific subgroups (containing at least 20% of subjects in the sample) most affected by the interaction.^20^ We then extracted the two-way chemical combinations and their thresholds indicated to interact by the rh-SiRF algorithm to define ‘low-low’, ‘low-high’, ‘high-low’ and ‘high-high’ exposure groups (based on the identified chemical interactions by rh-SiRF), and performed a downstream linear regression to obtain the effect estimates for the two-way chemical interaction subgroup combinations.

A quantile-based threshold-finding algorithm is implemented that transforms the combinations of exposures into indicators or “chemical-cliques” using their thresholds.^20^ For example, considering a combination of chemicals A and B, which was converted into a 2^nd^-order chemical-clique “low A/high B”, as an indicator function. Here, chemical-clique “low A/high B” implies lower concentrations of metals A (based on certain thresholds) and a higher concentration of metal B in the sample. This 2^nd^-order chemical-clique denotes an underlying sub-sample (or subgroup) satisfying the conditions of the clique. The final chemical-cliques were selected based on a frequency above 2.5% within all the cliques detected. All cliques had also at least a 20% sample prevalence (within the cohort) to address overfitting.

**Supplementary Methods 8. Handling Missing Data and Additional Sensitivity Analyses Methods.**

Missing values imputed in analyses by implementing the *missForest* package using a random forest with ten maximum iterations and 100 trees. Observations with missing values included participants with missing metals (0.9%-1.0%), pesticides (46%-50%), air pollutants (7.3%-11%), puberty (4.4%), and FA (7.8%-8.5%) (**Tables 1-2**, **Table S3**). We also conducted several secondary analyses: 1) we examined potential effect modification (stratified analyses) by sex, puberty status, and overweight/obesity on liver injury, given that sex-, obesity-, and puberty-dependent associations between MDCs and metabolic outcomes have been proposed previously.^23^ Effect modification by these factors was examined using the cross-product term between the dichotomized effect modifier and the BWQS weighted index mean. 2) We also repeated the MDC-ALT analyses using a cutoff of ALT≥25 to define liver injury in children.^5^ 3) Additional MDC-liver injury models in children were further adjusted for other important lifestyle predictors of liver injury and/or metabolic syndrome identified prior in our data, such as sugar-sweetened beverages (SSB) and sedentary time. 4) Given the high number of missing data in organophosphate pesticide measurements, we also performed sensitivity analyses restricted to mother-child pairs with measured organophosphate pesticide data.

| **Table S1. Baseline Participant Characteristics in PROGRESS (n=948 Mothers) and Subset with Liver Enzyme Data (n=234 Mothers).^a^** | | |
| --- | --- | --- |
| **Variable** | **N = 948** | **N = 234** |
| **Mother Age at Partum (in years)** | 28.0 (5.49) [18.0, 44.0] | 28.1 (5.39) [19.0, 44.0] |
| **AMAI SES Index** |  |  |
| Low | 486 (51.3%) | 123 (53.4%) |
| Medium | 356 (37.6%) | 86 (36.8%) |
| High | 106 (11.2%) | 23 (9.8%) |
| **Any Smoking during Pregnancy** |  |  |
| No Smoking (Any) during Pregnancy | 596 (62.9%) | 160 (68.4%) |
| Smoking (Any) during Pregnancy | 352 (37.1%) | 74 (31.6%) |
| **Parity at Baseline** |  |  |
| 1 pregnancy | 355 (37.4%) | 94 (40.2%) |
| 2 pregnancies | 339 (35.8%) | 80 (34.2%) |
| 3+ pregnancies | 254 (26.8%) | 60 (25.6%) |
| **Pre-pregnancy BMI** | 26.3 (4.16) [17.1, 43.5] | 26.5 (4.27) [18.6, 40.5] |
| **Alcohol during Pregnancy** |  |  |
| No Drinking during Pregnancy | 841 (88.7%) | 193 (82.5%) |
| Drinking during Pregnancy | 107 (11.3%) | 41 (17.5%) |
| ^a^ Mean (SD) [Range: min, max]; n (%). |  |  |

| **Table S2. PROGRESS Liver Enzyme Distributions Among Two Pesticide Group Subsets.** | | |
| --- | --- | --- |
| **Liver Enzyme** | **Mean (SD) [Range]**  **(Pesticide Group 1) ^a^** | **Mean (SD) [Range]**  **(Pesticide Group 2) ^a^** |
| **Mothers** | **N = 81** | **N = 36** |
| **ALT** | 15.0 (9.88) [3.20, 54.4] | 14.7 (8.52) [3.20, 37.4] |
| **Ln(ALT)** | 2.61 (0.57) [1.44, 4.01] ^b^ | 2.62 (0.54) [1.44, 3.65] |
| **AST** | 18.6 (7.67) [4.80, 52.3] | 17.4 (6.44) [8.90, 40.6] |
| **Ln(AST)** | 2.84 (0.41) [1.57, 3.96] ^b^ | 2.80 (0.33) [2.19, 3.70] |
| **Children** | **N = 68** | **N = 36** |
| **ALT** | 13.7 (11.5) [3.00, 79.8] | 13.7 (10.2) [4.50, 52.1] |
| **Ln(ALT)** | 2.51 (0.55) [1.39, 4.39] | 2.53 (0.53) [1.70, 3.97] |
| **AST** | 25.3 (8.02) [14.8, 66.6] | 24.2 (8.01) [13.8, 56.3] |
| **Ln(AST)** | 3.19 (0.28) [2.69, 4.20] | 3.14 (0.29) [2.62, 4.03] |
| ^a^ Mean (SD) [Range]. Group 1 refers to phase 1 pesticides whereas group 2 refers to phase 2 pesticides (**Supplementary Methods 2**).  ^b^ 2 outliers excluded (shown as missing values). | | |

| **Table S3. Chemical Exposure Distributions for Analyses in PROGRESS between 2007-2011. ^a^** | | |
| --- | --- | --- |
| **Variable** | **Children (n = 205)** | **Mothers (n = 234)** |
| ***Air Pollutants*** |  |  |
| **Particulate Matter 2.5 (PM_2.5_ in μg/m^3^) ^b^** | 20.0 (4.19) [16.0, 29.5] | 20.2 (4.06) [16.0, 29.5] |
| **Nitrogen Dioxide (NO_2_ in μg/m^3^) ^c^** | 31.6 (6.80) [17.7, 47.5] | 31.7 (6.99) [17.7, 47.5] |
| ***Non-Imputed Air Pollutants*** |  |  |
| **Particulate Matter 2.5 (PM_2.5_ in μg/m^3^)** | 19.9 (4.37) [16.0, 29.5] | 20.1 (4.60) [16.0, 29.5] |
| **Nitrogen Dioxide (NO_2_ in μg/m^3^)** | 31.1 (7.76) [17.6, 48.6] | 31.4 (8.42) [17.6, 47.5] |
| ***Pesticide Metabolites ^d^*** |  |  |
| **Diethylthiophosphate (DETP in ng/mL)** | 1.02 (0.99) [0.11, 18.1] | 0.72 (0.53) [0.11, 9.99] |
| **Dimethyldithiophosphate (DMDTP in ng/mL)** | 0.66 (0.63) [0.11, 11.4] | 0.67 (0.66) [0.11, 11.4] |
| **Dimethylthiophosphate (DMTP in ng/mL)** | 5.06 (6.43) [0.21, 72.9] | 4.88 (5.31) [0.21, 72.9] |
| **Nitrophenol (PNP in ng/mL)** | 2.58 (1.96) [0.43, 36.9] | 2.08 (2.08) [0.43, 36.9] |
| **3,5,6-Trichloro-2-Pyridinol (TCP in ng/mL)** | 2.06 (1.92) [0.13, 36.5] | 1.83 (1.65) [0.13, 36.5] |
| ***Non-Imputed and Unadjusted Pesticide Metabolites ^e^*** |  |  |
| **Diethylthiophosphate (DETP in ng/mL)** | 0.45 (0.44) [0.14, 14.1] | 0.43 (0.48) [0.14, 14.1] |
| **Dimethyldithiophosphate (DMDTP in ng/mL)** | 0.35 (0.38) [0.14, 6.35] | 0.35 (0.39) [0.14, 6.35] |
| **Dimethylthiophosphate (DMTP in ng/mL)** | 3.11 (5.91) [0.35, 54.2] | 3.29 (5.56) [0.35, 54.2] |
| **Nitrophenol (PNP in ng/mL)** | 1.81 (2.03) [0.26, 32.0] | 1.88 (2.00) [0.26, 32.0] |
| **3,5,6-Trichloro-2-Pyridinol (TCP in ng/mL)** | 1.34 (1.56) [0.18, 28.3] | 1.30 (1.65) [0.18, 28.3] |
| ***Non-Imputed and Unadjusted Pesticide Metabolites 60%<LOD ^e^*** |  |  |
| **Diethyldithiophosphate (DEDTP in ng/mL)** | 0.07 (0.28) [0.07, 7.11] | 0.07 (0.28) [0.07, 7.11] |
| **Malathion dicarboxylic acid (MDA in ng/mL)** | 0.14 (0.21) [0.14, 2.50] | 0.14 (0.21) [0.14, 2.50] |
| ***Essential Metals or Trace Elements*** |  |  |
| **Selenium (Se in μg/L) ^f^** | 236 (28.2) [183, 363] | 236 (30.3) [183, 458] |
| **Zinc (Zn in μg/L) ^f^** | 5,853 (1,076) [4,098, 9,317] | 5,950 (1,120) [4,098, 10,232] |
| **Cobalt (Co in** **μg/L) ^f^** | 0.24 (0.12) [0.07, 3.45] | 0.23 (0.12) [0.07, 3.45] |
| **Copper (Cu in μg/L) ^f^** | 1,489 (216) [1,089, 2,576] | 1,494 (219) [1,089, 2,576] |
| **Magnesium (Mg in μg/L) ^g^** | 68,436 (47,168) [15,156, 583,564] | 70,857 (49,337) [18,393, 583,564] |
| **Manganese (Mn in μg/L) ^f^** | 14.9 (6.33) [5.60, 40.1] | 14.9 (6.34) [5.60, 40.1] |
| **Molybdenum (Mo in μg/L) ^g^** | 44.2 (32.5) [8.63, 336] | 45.0 (29.4) [8.63, 336] |
| ***Non-Essential Elements or Metals*** |  |  |
| **Lead (Pb in μg/L) ^f^** | 28.9 (29.2) [8.25, 170] | 28.8 (28.9) [8.25, 170] |
| **Arsenic (As in μg/L) ^g^** | 15.4 (13.0) [3.25, 297] | 15.6 (13.0) [3.25, 297] |
| **Chromium (Cr in μg/L) ^g^** | 0.68 (0.53) [0.33, 5.08] | 0.69 (0.56) [0.33, 5.39] |
| **Cesium (Cs in μg/L) ^g^** | 7.86 (3.81) [1.88, 55.6] | 7.97 (4.15) [1.88, 55.6] |
| **Nickel (Ni in μg/L) ^g^** | 3.49 (2.41) [1.44, 140] | 3.49 (2.26) [1.12, 140] |
| **Antimony (Sb in μg/L) ^g^** | 0.13 (0.09) [0.03, 1.39] | 0.13 (0.09) [0.03, 1.39] |
| **Aluminum (Al in μg/L) ^g^** | 39.6 (30.0) [12.4, 720] | 38.9 (26.5) [12.4, 720] |
| **Barium (Ba in μg/L) ^g^** | 4.45 (4.23) [0.57, 40.7] | 4.44 (3.96) [0.19, 38.8] |
| **Mercury (Hg in μg/L) ^g^** | 0.99 (1.23) [0.25, 23.8] | 0.98 (1.07) [0.25, 23.8] |
| **Tin (Sn in μg/L) ^g^** | 1.58 (2.80) [0.16, 53.6] | 1.53 (2.49) [0.25, 33.4] |
| **Strontium (Sr in μg/L) ^g^** | 165 (120) [18.3, 879] | 173 (116) [31.6, 879] |
| **Thallium (Tl in μg/L) ^g^** | 0.37 (0.21) [0.08, 2.60] | 0.37 (0.21) [0.08, 2.60] |
| **Vanadium (V in μg/L) ^g^** | 0.51 (0.37) [0.15, 4.64] | 0.53 (0.36) [0.12, 4.64] |
| **Cadmium (Cd in μg/L) ^g^** | 0.21 (0.19) [0.05, 4.45] | 0.21 (0.18) [0.04, 4.45] |
| ***Non-Imputed and Unadjusted Elements or Metals*** |  |  |
| **Selenium (Se in μg/L) ^h^** | 236 (29) [183, 363] | 236 (30.5) [183, 458] |
| **Zinc (Zn in μg/L) ^h^** | 5,879 (1,085) [4,098, 9,317] | 5,940 (1,125) [4,098, 10,232] |
| **Cobalt (Co in μg/L) ^h^** | 0.24 (0.12) [0.07, 3.45] | 0.23 (0.12) [0.07, 3.45] |
| **Copper (Cu in μg/L) ^h^** | 1,489 (219) [1,089, 2,576] | 1,494 (221) [1,089, 2,576] |
| **Manganese (Mn in μg/L) ^h^** | 14.9 (6.34) [5.60, 40.1] | 14.8 (6.43) [5.60, 40.1] |
| **Lead (Pb in μg/L) ^h^** | 28.8 (29.4) [8.25, 170] | 28.5 (29.2) [8.25, 170] |
| **Magnesium (Mg in μg/L) ^e^** | 63,450 (43,350) [15,145, 203,000] | 69,100 (47,138) [15,145, 203,000] |
| **Molybdenum (Mo in μg/L) ^e^** | 42.4 (29.2) [8.13, 297] | 43.4 (28.8) [8.13, 297] |
| **Arsenic (As in μg/L) ^e^** | 14.3 (11.2) [2.62, 279] | 14.9 (10.6) [2.62, 279] |
| **Chromium (Cr in μg/L) ^e^** | 0.58 (0.00) [0.58, 3.76] | 0.58 (0.00) [0.58, 4.08] |
| **Cesium (Cs in μg/L) ^e^** | 7.89 (4.39) [2.06, 25.9] | 8.11 (4.24) [2.06, 25.9] |
| **Nickel (Ni in μg/L) ^e^** | 3.24 (1.48) [1.62, 55.0] | 3.30 (1.54) [1.62, 55.0] |
| **Antimony (Sb in μg/L) ^e^** | 0.13 (0.06) [0.04, 0.70] | 0.13 (0.07) [0.04, 0.70] |
| **Aluminum (Al in μg/L) ^e^** | 34.9 (20.3) [16.7, 295] | 33.7 (19.2) [16.7, 295] |
| **Barium (Ba in μg/L) ^e^** | 3.67 (3.17) [0.57, 41.8] | 3.70 (3.42) [0.29, 41.8] |
| **Mercury (Hg in μg/L) ^e^** | 0.96 (1.09) [0.21, 30.7] | 0.98 (1.05) [0.21, 30.7] |
| **Tin (Sn in μg/L) ^e^** | 1.46 (2.37) [0.21, 20.3] | 1.41 (2.22) [0.21, 30.2] |
| **Strontium (Sr in μg/L) ^e^** | 150 (88.1) [26.0, 522] | 155 (100) [33.3, 637] |
| **Thallium (Tl in μg/L) ^e^** | 0.36 (0.24) [0.09, 1.14] | 0.38 (0.23) [0.09, 1.29] |
| **Vanadium (V in μg/L) ^e^** | 0.44 (0.26) [0.20, 2.38] | 0.47 (0.27) [0.18, 2.65] |
| **Cadmium (Cd in μg/L) ^e^** | 0.19 (0.16) [0.05, 3.69] | 0.20 (0.16) [0.05, 3.69] |
| ***High-Molecular-Weight-Phthalate (HMWP) Metabolites ^g^*** |  |  |
| **Mono-2-ethylhexyl phthalate (MEHP in ng/mL)** | 6.17 (6.99) [0.52, 113] | 6.26 (7.01) [0.52, 113] |
| **Mono-2-ethyl-5-oxohexyl phthalate (MEOHP in ng/mL)** | 21.2 (21.3) [1.89, 288] | 21.5 (22.5) [1.89, 288] |
| **Mono-2-ethyl-5-hydroxyhexyl phthalate (MEHHP in ng/mL)** | 22.6 (24.8) [1.82, 317] | 22.6 (26.5) [1.82, 317] |
| **Mono-2-ethyl-5-carboxypentyl phthalate (MECPP in ng/mL)** | 46.2 (48.8) [5.87, 779] | 46.4 (50.7) [5.87, 779] |
| **Mono-2-ethyl-5-carboxypentyl terephthalate (MECPTP in** **ng/mL)** | 3.22 (4.08) [0.20, 80.7] | 3.23 (3.93) [0.33, 80.7] |
| **Monooxononyl phthalate (MONP in ng/mL)** | 1.65 (1.82) [0.16, 42.0] | 1.64 (1.76) [0.16, 42.0] |
| **Mono carboxyisooctyl phthalate (MCOP in ng/mL)** | 5.99 (4.84) [0.73, 60.3] | 5.06 (4.44) [0.69, 60.3] |
| **Mono carboxyisononyl phthalate (MCNP in ng/mL)** | 1.06 (0.85) [0.20, 26.9] | 1.07 (0.80) [0.22, 26.9] |
| **Mono-3-carboxypropyl phthalate (MCPP in ng/mL)** | 1.51 (1.37) [0.23, 23.4] | 1.63 (1.47) [0.31, 23.4] |
| **Monobenzyl phthalate (MBzP in ng/mL)** | 5.31 (7.34) [0.23, 83.9] | 5.99 (8.56) [0.23, 83.9] |
| ***Unadjusted HMWP Metabolites ^e^*** |  |  |
| **Mono-2-ethylhexyl phthalate (MEHP in ng/mL)** | 5.60 (6.10) [0.57, 60.4] | 5.88 (5.94) [0.57, 60.4] |
| **Mono-2-ethyl-5-oxohexyl phthalate (MEOHP in ng/mL)** | 19.7 (22.2) [2.10, 232] | 20.8 (23.6) [2.10, 232] |
| **Mono-2-ethyl-5-hydroxyhexyl phthalate (MEHHP in ng/mL)** | 22.8 (22.8) [1.70, 254] | 23.1 (25.3) [1.70, 254] |
| **Mono-2-ethyl-5-carboxypentyl phthalate (MECPP in ng/mL)** | 41.5 (45.1) [4.60, 544] | 43.5 (50.2) [4.60, 544] |
| **Mono-2-ethyl-5-carboxypentyl terephthalate (MECPTP in ng/mL)** | 2.85 (3.25) [0.14, 63.2] | 3.00 (3.18) [0.14, 63.2] |
| **Monooxononyl phthalate (MONP in ng/mL)** | 1.50 (1.60) [0.28, 40.6] | 1.55 (1.53) [0.28, 40.6] |
| **Mono carboxyisooctyl phthalate (MCOP in ng/mL)** | 4.50 (3.75) [0.70, 58.4] | 4.58 (4.03) [0.70, 58.4] |
| **Mono carboxyisononyl phthalate (MCNP in ng/mL)** | 0.97 (0.75) [0.70, 22.2] | 1.00 (0.75) [0.20, 22.2] |
| **Mono-3-carboxypropyl phthalate (MCPP in ng/mL)** | 1.35 (1.36) [0.28, 12.4] | 1.50 (1.30) [0.28, 13.3] |
| **Monobenzyl phthalate (MBzP in ng/mL)** | 5.10 (6.05) [0.21, 66.0] | 5.73 (7.78) [0.21, 66.0] |
| ***Low-Molecular-Weight Phthalate (LMWP) Metabolites ^g^*** |  |  |
| **Mono-hydroxyisobutyl phthalate (MHiBP in ng/mL)** | 3.83 (4.71) [0.35, 31.2] | 4.05 (4.63) [0.35, 41.4] |
| **Mono-isobutyl phthalate (MiBP in ng/mL)** | 9.15 (11.4) [0.58, 97.1] | 9.31 (12.3) [0.84, 97.2] |
| **Mono-n-butyl phthalate (MBP in ng/mL)** | 85.2 (97.6) [4.28, 2,439] | 93.2 (112.3) [4.82, 2,439] |
| **Mono-hydroxybutyl phthalate (MHBP in ng/mL)** | 7.76 (10.5) [0.46, 269] | 8.38 (10.6) [0.52, 269] |
| **Monoethyl phthalate (MEP in ng/mL)** | 125 (278) [6.25, 3,493] | 144 (293) [6.25, 4,941] |
| ***Unadjusted LMWP Metabolites ^e^*** |  |  |
| **Mono-hydroxyisobutyl phthalate (MHiBP in ng/mL)** | 3.35 (4.25) [0.28, 30.0] | 3.63 (4.58) [0.28, 49.8] |
| **Mono-isobutyl phthalate (MiBP in ng/mL)** | 8.70 (11.2) [0.78, 66.3] | 9.73 (11.5) [0.78, 123] |
| **Mono-n-butyl phthalate (MBP in ng/mL)** | 71.3 (104) [2.40, 1,300] | 83.9 (110) [2.40, 1,300] |
| **Mono-hydroxybutyl phthalate (MHBP in ng/mL)** | 7.10 (9.15) [0.28, 141] | 7.95 (8.88) [0.28, 172] |
| **Monoethyl phthalate (MEP in ng/mL)** | 109 (257) [5.50, 2,640] | 132 (271) [5.50, 3,192] |
| ^a^ Median (IQR) [Range]. Distributions showing concentrations of chemicals already imputed for missing values (using random forest with 10 maximum iterations and 100 trees) unless specified. All chemicals shown with concentrations below LOD were imputed with LOD/√2. Only chemicals with at least 60% of observations >LOD were included in subsequent statistical analyses. The distributions for chemical exposures are shown before log_2_-transformation in this table. For chemicals measured more than once in pregnancy, averaged concentrations across trimesters are shown (i.e. average of all three trimester values for air pollutants, and average of trimesters 2 and 3 values for metal/metalloids and phthalates). | | |
| ^b^ n=16 missing values in children and n=25 missing values in mothers (imputed using random forest). | | |
| ^c^ n=15 missing values in children and n=24 missing values in mothers (imputed using random forest). | | |
| ^d^ Concentrations measured in urine and corrected for specific gravity. There were n=101 missing values in children and n=117 missing values in mothers (imputed using random forest). | | |
| ^e^ Concentrations measured in urine and not corrected for specific gravity. | | |
| ^f^ Concentrations measured in blood. There were n=2 missing values (imputed using random forest). | | |
| ^g^ Concentrations measured in urine and corrected for specific gravity. | | |
| ^h^ Measured in blood. | | |

| **Table S4. Characteristics and Outcomes in PROGRESS Children.** | |
| --- | --- |
| **Variable** | **N = 205 ^a^** |
| **Maternal Age at Partum (in years)** | 28.3 (5.53) [19.0, 44.0] |
| **SES Index** |  |
| Low | 113 (55.1%) |
| Medium | 74 (36.1%) |
| High | 18 (8.8%) |
| **Smoking Exposure (Passive/Active) during Pregnancy** |  |
| No | 142 (69.3%) |
| Yes | 63 (30.7%) |
| **Parity at Baseline (including index pregnancy)** |  |
| 1 pregnancy | 78 (38.0%) |
| 2 pregnancies | 68 (33.2%) |
| 3+ pregnancies | 59 (28.8%) |
| **Maternal Pre-pregnancy Body Mass Index (BMI in kg/m^2^)** | 26.6 (4.22) [18.6, 40.5] |
| **Alcohol during Pregnancy ^b^** |  |
| No | 165 (80.5%) |
| Yes | 40 (19.5%) |
| **Maternal Pregnancy FA Intake (µg/day) ^c^** | 503 (182) [7.50, 1,350] |
| **Maternal Pregnancy FA Intake ≥400 µg/day ^c^** | 181 (88.3%) |
| **Maternal Pregnancy FA Intake ≥600 µg/day ^c^** | 38 (18.5%) |
| **Child Exact Age at ~9-Year Examination** | 9.36 (0.86) [8.08, 12.1] |
| **Sex** |  |
| Female | 99 (48.3%) |
| Male | 106 (51.7%) |
| **Puberty ^d^** |  |
| Pre-puberty (Tanner stage=1) | 43 (21.0%) |
| Puberty (Tanner stages=2-5) | 162 (79.0%) |
| **Child zBMI at Age ~9 Years ^e^** | 0.91 (1.27) [-2.39, 3.54] |
| **Child Overweight (WHO reference) at ~9 Years ^e^** | 92 (45.1%) |
| **Alanine Transaminase (ALT in U/L) at ~9 Years^f^** | 14.0 (10.7) [3.00, 79.8] |
| **Aspartate Aminotransferase (AST in U/L) at ~9 Years^g^** | 25.0 (8.44) [9.20, 74.1] |
| **Gamma-glutamyltransferase (GGT in U/L) at ~9 Years^h^** | 13.2 (5.30) [4.10, 40.0] |
| **ALT-elevation (≥25.3 U/L) ^f,i^** | 20 (9.9%) |
| **ALT-elevation (≥25 U/L) ^f^** | 22 (10.7%) |
| **AST-elevation (≥32.5 U/L) ^g,i^** | 21 (10.8%) |
| **Pediatric NAFLD fibrosis score (PNFS) ^j^** | 2.57 (1.32) [0.77, 10.5] |
| **PNFS (≥8) ^j,k^** | 1 (0.5%) |
| ^a^ Mean (SD) [Range: min, max]; n (%). The distributions for the following outcomes are shown in the original scale (not ln-transformed): ALT, AST, and GGT. | |
| ^b^ Note heavy alcohol drinkers during pregnancy were excluded from PROGRESS cohort enrollment. n=19 imputed missing values using random forest with 10 maximum iterations and 100 trees. | |
| ^c^ n=16 imputed missing values using random forest with 10 maximum iterations and 100 trees. | |
| ^d^ n=9 imputed missing values using random forest with 10 maximum iterations and 100 trees. | |
| ^e^ n=1 missing value (effect modifier) excluded for effect modification analyses. Childhood BMI during the year ~9 examination was based on body mass index (BMI) z-scores (World Health Organization, WHO, ages >5) where children 1 standard deviation (SD) above the WHO Growth Reference median were considered overweight. | |
| ^f^ n=2 outliers excluded (based on Rosner's generalized extreme Studentized deviate test). | |
| ^g^ n=1 outlier excluded (based on Rosner's generalized extreme Studentized deviate test). | |
| ^h^ n=3 outliers excluded (based on Rosner's generalized extreme Studentized deviate test). | |
| ^i^ Above 90^th^-percentile in the levels of liver enzymes (ALT, AST) in PROGRESS. | |
| ^j^ n=16 missing values. Index constructed based on the following formula: z=1.1+0.34*√ALT + 0.002*(Alkaline phosphatase) - 1.1*log(platelets) - 0.02*GGT. PFNS probability= [e^z^ / (1+e^z^)]*100.  ^k^ At risk of fibrosis.^24^ | |

| **Table S5. Liver Enzyme Correlations PROGRESS. ^a^** | | | |
| --- | --- | --- | --- |
| **Liver Enzyme** | **n** |  | **Spearman** |
| **ALT** | 189 |  | ρ=0.18 (p-value=0.011*) |
| **AST** | 190 |  | ρ=0.23 (p-value=0.001*) |

**P*-value<0.05

^a^ Using the non-ln-transformed values of liver enzymes. Overlapping population in which both mother-child (pair) had liver enzymes measured.

**Figure S1. Chemical Distributions and Correlations in PROGRESS Participants. ^a^**

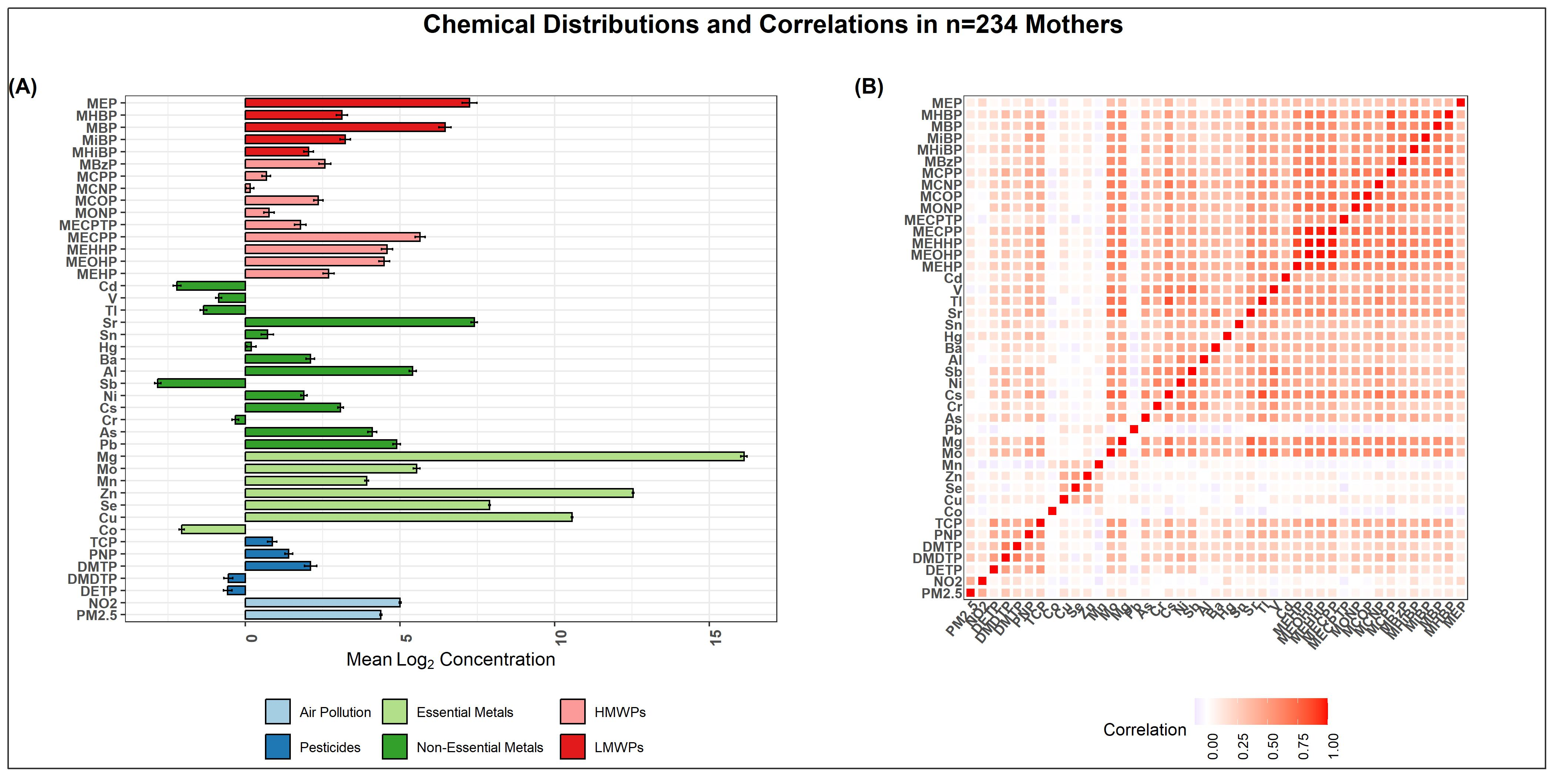

^a^ Metabolism-Disrupting Chemical (MDC) concentrations in mothers during pregnancy are shown in graph (A). Error bars indicate 2*SE log (base=2) concentration. Error bars are color-coded as follows: light blue denotes air pollution, dark blue denotes pesticides, light green denotes essential metals or trace elements, dark green denotes non-essential elements or metals, light red denotes high-molecular-weight phthalates, and dark red denotes low-molecular-weight phthalates. Correlation plots are shown in heatmap (B).

| **Table S6. Estimated Posterior Weights of HMWP-Mixture Group on ALT using BWQS Models in PROGRESS Children. ^a^** | | | |
| --- | --- | --- | --- |
| HMWP | **ALT** (Continuous) | HMWP | **ALT Elevation**  (Dichotomized ALT ≥90th percentile Cutoff) |
| MECPP | 0.10918 | MCOP | 0.11412 |
| MCNP | 0.10515 | MECPTP | 0.10846 |
| MEOHP | 0.10490 | MEHP | 0.10413 |
| MONP | 0.10313 | MCNP | 0.10367 |
| MCOP | 0.10288 | MECPP | 0.10084 |
| MEHHP | 0.10234 | MEHHP | 0.09679 |
| MEHP | 0.09968 | MONP | 0.09462 |
| MBzP | 0.09339 | MBzP | 0.09366 |
| MECPTP | 0.09149 | MCPP | 0.09261 |
| MCPP | 0.08785 | MEOHP | 0.09110 |
| ^a^ Ordered by high to low estimated posterior weight. | | | |

| **Table S7. Estimated Posterior Weights of Phthalate-Mixture Group on Continuous AST using BWQS Models in PROGRESS Children. ^a^** | |
| --- | --- |
| **HMWP** | **AST** |
| MCOP | 0.11105 |
| MONP | 0.10698 |
| MBzP | 0.10492 |
| MCNP | 0.10225 |
| MECPTP | 0.09969 |
| MCPP | 0.09873 |
| MEHP | 0.09700 |
| MECPP | 0.09540 |
| MEHHP | 0.09459 |
| MEOHP | 0.08940 |
| **LMWP** | **AST** |
| MiBP | 0.23108 |
| MBP | 0.22375 |
| MHiBP | 0.22273 |
| MHBP | 0.19422 |
| MEP | 0.12822 |
| ^a^ Ordered by high to low estimated posterior weight. | |

**Figure S2. Global (Overall) BWQS with All Metabolism-Disrupting Chemicals in Relation to Liver Injury in PROGRESS Children.**

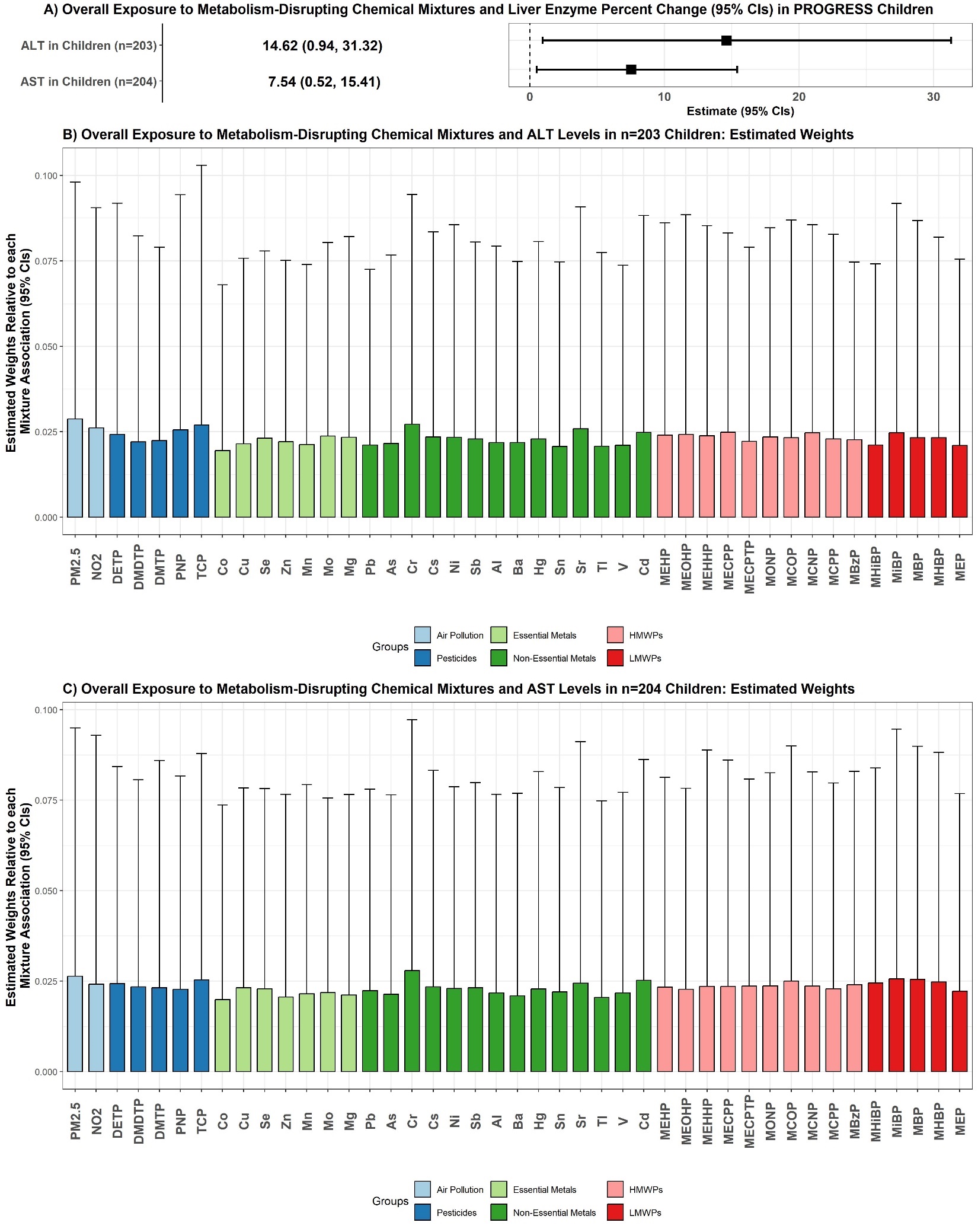

^a^ Forest plots of overall mixtures (including all MDCs) on liver enzymes ALT and AST (A) in PROGRESS children using covariate-adjusted Bayesian Weighted Quantile Sum (BWQS) regression. In forest plot A, squares represent effect estimates and whiskers represent 95% confidence intervals. Ln-transformed continuous liver enzyme estimates were back-transformed from the log-scale by exponentiating the value, subtracting 1, and multiplying by a 100. Estimated posterior weights are also shown (B, C) for models (ALT and AST global BWQS in children), representing the estimated contribution of each chemical exposure to the overall group association. Posterior weights are color-coded as follows: light blue denotes air pollution, dark blue denotes pesticides, light green denotes essential metals or trace elements, dark green denotes non-essential elements or metals, light red denotes high-molecular-weight phthalates, and dark red denotes low-molecular-weight phthalates. All models in children were adjusted for child age at follow-up year ~9, continuous maternal pre-pregnancy BMI, SES, sex, maternal parity, maternal smoking status and alcohol intake during pregnancy, and age at parturition.

| **Table S8. Estimated Posterior Weights of Chemical-Mixture Groups on Liver Injury using Overall BWQS Models in PROGRESS Children. ^a^** | | | | |
| --- | --- | --- | --- | --- |
| MDC | **ALT** |  | MDC | **AST** |
| PM_2.5_ | 0.02880 |  | Cr | 0.02793 |
| Cr | 0.02720 |  | PM_2.5_ | 0.02636 |
| TCP | 0.02700 |  | MiBP | 0.02560 |
| NO_2_ | 0.02613 |  | MBP | 0.02550 |
| Sr | 0.02588 |  | TCP | 0.02535 |
| PNP | 0.02561 |  | Cd | 0.02524 |
| MECPP | 0.02489 |  | MCOP | 0.02506 |
| Cd | 0.02481 |  | MHBP | 0.02476 |
| MCNP | 0.02476 |  | MHiBP | 0.02448 |
| MiBP | 0.02475 |  | Sr | 0.02445 |
| MEOHP | 0.02426 |  | DETP | 0.02428 |
| DETP | 0.02424 |  | NO_2_ | 0.02413 |
| MEHP | 0.02402 |  | MBzP | 0.02401 |
| MEHHP | 0.02385 |  | MONP | 0.02367 |
| Mo | 0.02375 |  | MECPTP | 0.02365 |
| MONP | 0.02345 |  | MCNP | 0.02361 |
| Cs | 0.02344 |  | MECPP | 0.02357 |
| Mg | 0.02339 |  | MEHHP | 0.02356 |
| Ni | 0.02337 |  | DMDTP | 0.02340 |
| MCOP | 0.02324 |  | Cs | 0.02340 |
| MHBP | 0.02324 |  | MEHP | 0.02333 |
| MBP | 0.02322 |  | Cu | 0.02317 |
| Se | 0.02310 |  | Sb | 0.02317 |
| MCPP | 0.02293 |  | DMTP | 0.02314 |
| Sb | 0.02288 |  | Ni | 0.02294 |
| Hg | 0.02287 |  | MCPP | 0.02292 |
| MBzP | 0.02270 |  | Se | 0.02291 |
| DMTP | 0.02242 |  | Hg | 0.02284 |
| MECPTP | 0.02221 |  | MEOHP | 0.02276 |
| Zn | 0.02214 |  | PNP | 0.02273 |
| DMDTP | 0.02208 |  | Pb | 0.02234 |
| Al | 0.02182 |  | MEP | 0.02225 |
| Ba | 0.02181 |  | Sn | 0.02201 |
| As | 0.02155 |  | Mo | 0.02186 |
| Cu | 0.02152 |  | Al | 0.02178 |
| Mn | 0.02130 |  | V | 0.02177 |
| Pb | 0.02113 |  | Mn | 0.02151 |
| MHiBP | 0.02113 |  | As | 0.02133 |
| V | 0.02109 |  | Mg | 0.02119 |
| MEP | 0.02098 |  | Ba | 0.02098 |
| Tl | 0.02079 |  | Zn | 0.02064 |
| Sn | 0.02072 |  | Tl | 0.02049 |
| Co | 0.01952 |  | Co | 0.01990 |
| ^a^ Ordered by high to low estimated posterior weight. | |  |  |  |

**Figure S3. Associations Between Pregnancy MDC Mixtures and Liver Injury in PROGRESS Mothers (% Change [95% CIs] per Quartile Increases in MDC Mixtures). ^a^**

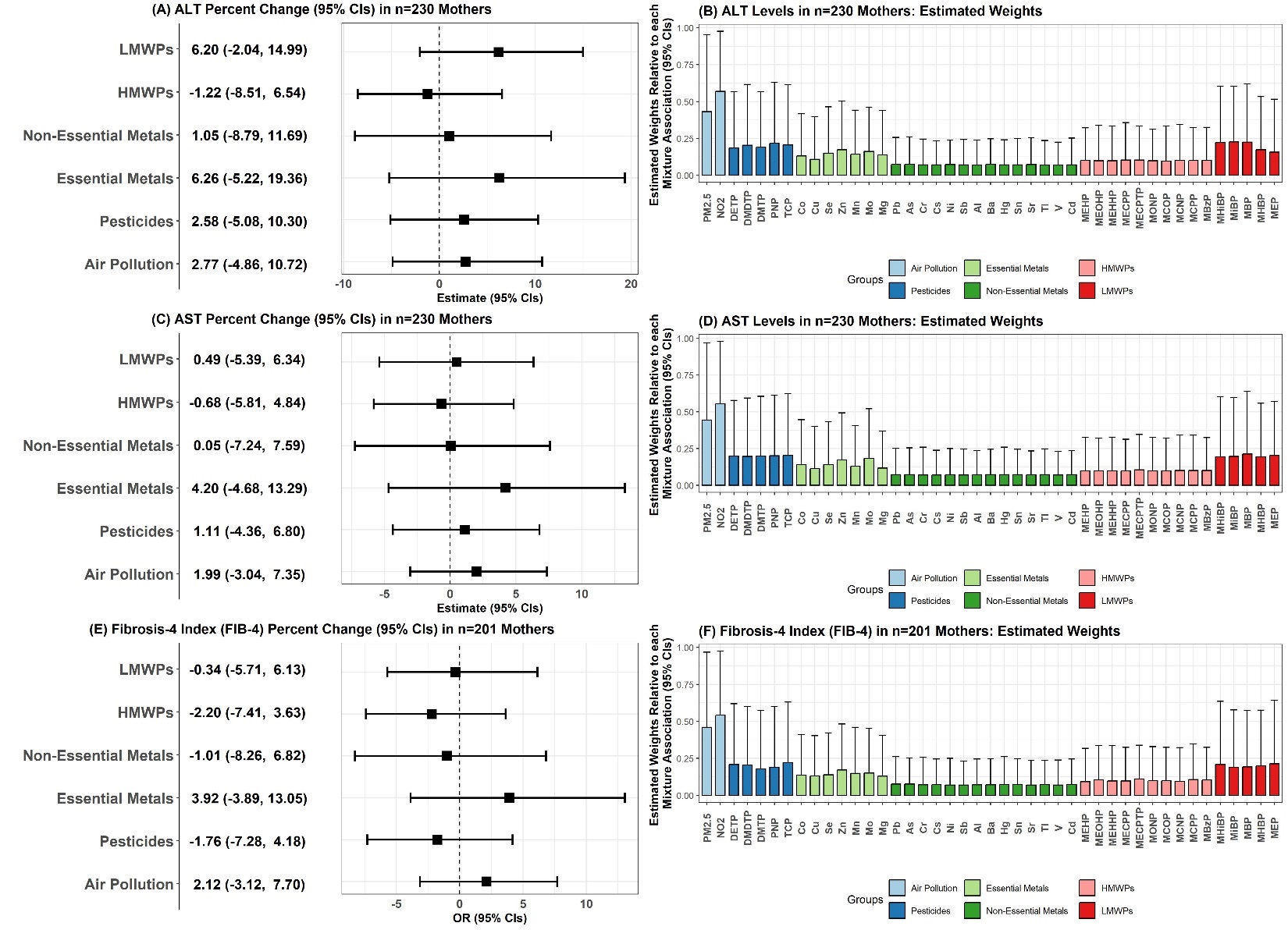

^a^ Forest plots and estimated posterior weights of MDC mixture groups and liver injury outcomes in PROGRESS mothers. On the left-hand side, forest plots (A, C, E) represent the effect estimates from the Bayesian Weighted Quantile Sum (BWQS) regression models, adjusted for continuous maternal pre-pregnancy BMI, SES, maternal parity, maternal smoking status and alcohol intake during pregnancy, and age at parturition. Each effect estimate represents different BWQS models conducted within each MDC group: squares represent effect estimates and whiskers represent 95% confidence intervals. Estimates were back-transformed from the log-scale by exponentiating the value, subtracting 1, and multiplying by a 100. Analyses with liver indexes were restricted to observations with available information only without missing items or components. On the right-hand side, graphs (B, D, F) represent the estimated weight or contribution of each chemical to the overall group mixture association (weighted index within each MDC group). Posterior weights are color-coded as follows: light blue denotes air pollutants, dark blue denotes organophosphate pesticide metabolites, light green denotes essential metals or trace elements, dark green denotes non-essential elements or metals, light red denotes high-molecular-weight phthalate metabolites, and dark red denotes low-molecular-weight phthalate metabolites.

**Figure S4. Global (Overall) BWQS with All Metabolism-Disrupting Chemicals in Relation to Liver Injury in PROGRESS Mothers.**

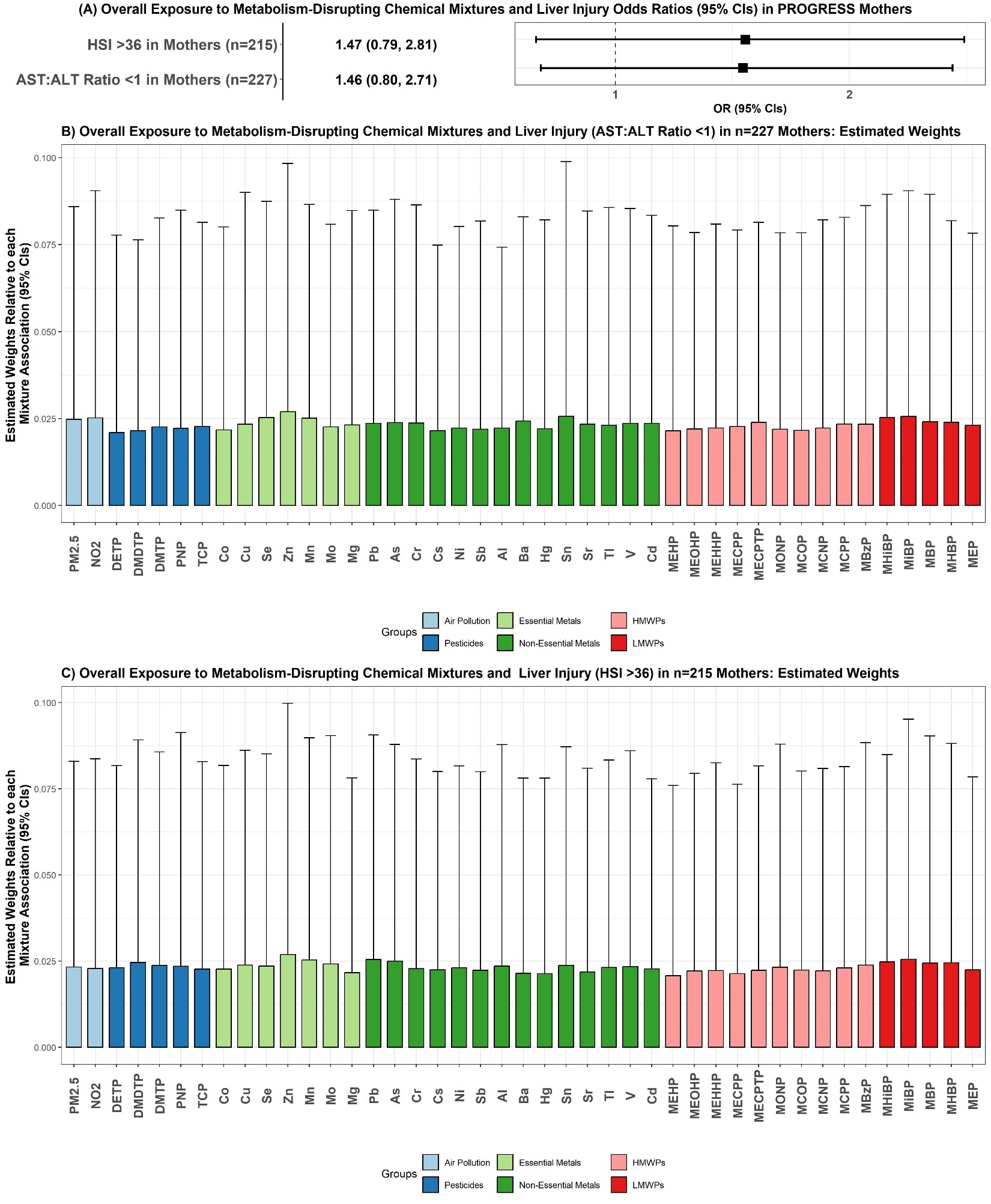

^a^ Forest plots of overall mixtures (including all MDCs) on liver injury outcomes (AST:ALT ratio <1 and HSI >36) (A) in PROGRESS mothers using covariate-adjusted Bayesian Weighted Quantile Sum (BWQS) regression. In forest plot A, squares represent effect estimates and whiskers represent 95% confidence intervals. Odds ratios are represented in the log_2_-scale for better visualization. Analyses with liver indexes were restricted to observations with available information only without missing items or components. Estimated posterior weights are also shown (B, C) for models, representing the estimated contribution of each chemical exposure to the overall group association. Posterior weights are color-coded as follows: light blue denotes air pollution, dark blue denotes pesticides, light green denotes essential metals or trace elements, dark green denotes non-essential elements or metals, light red denotes high-molecular-weight phthalate metabolites, and dark red denotes low-molecular-weight phthalate metabolites. Models in mothers were adjusted for continuous maternal pre-pregnancy BMI, SES, maternal parity, maternal smoking status and alcohol intake during pregnancy, and age at parturition.

**Figure S5. Rh-SiRF Analysis for Pre-selected Two-Order Chemical-Chemical Interactions in Relation to Liver Enzymes in PROGRESS Mothers. ^a^**

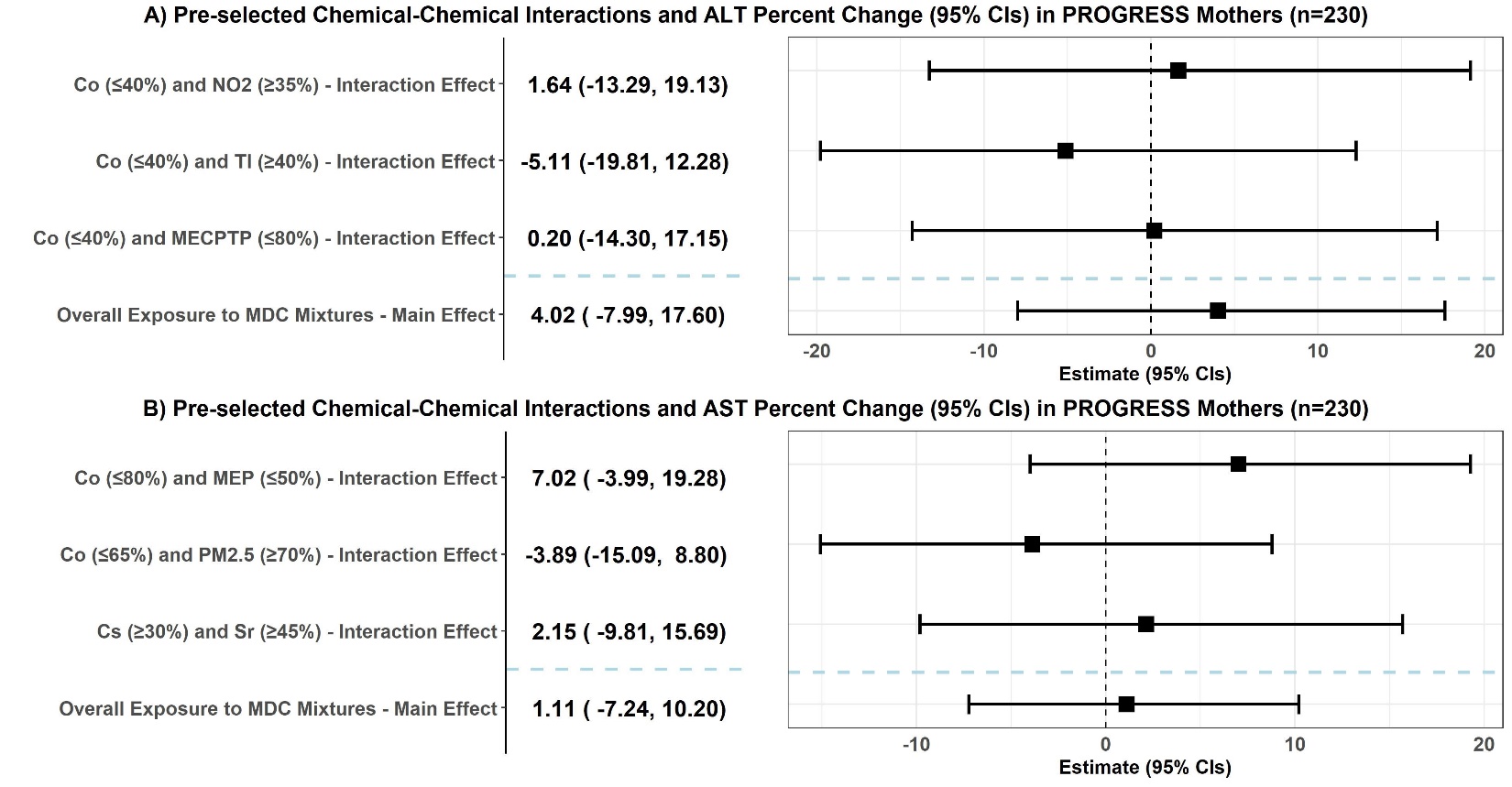

^a^ Results from repeated hold-out Signed Iterative Random Forest (rh-SiRF) for discovered interaction indicators in association with ln-transformed liver enzymes ALT and AST in mothers are shown in forest plots A and B, respectively. Estimates were back-transformed from the log-scale by exponentiating the value, subtracting 1, and multiplying by a 100. Linear models were fitted incorporating pre-selected interaction “cliques” (identified in children) as predictors of continuous liver enzyme endpoints in the mothers. In the top models, both the global BWQS chemical mixture (main effect) and the “clique” (interaction indicators) were introduced in the same model. For comparison, linear models using no chemical-chemical interactions as predictors with the global BWQS chemical mixture main effect were also visualized as the last model for each one of the forest plots. All models were adjusted for continuous maternal pre-pregnancy BMI, SES, maternal parity, maternal smoking status and alcohol intake during pregnancy, and age at parturition. The final chemical-cliques were selected based on a frequency above 2.5% within all the cliques detected for children applied in mothers. All cliques had also at least a 20% sample prevalence (within the cohort) to address overfitting.

| **Table S9. Effect Modification by Folic Acid Supplementation during Pregnancy in the Association Between MDC Mixtures with Liver Enzymes in PROGRESS Mothers.** | | | | | | | | | | |
| --- | --- | --- | --- | --- | --- | --- | --- | --- | --- | --- |
|  | |  | | |  | FA Supplementation Group ^a^ | | |  |  |
| Liver Outcome | | MDC Mixture | | |  | Percent Change (95% CIs) | | Percent Change (95% CIs) |  | *P*-for interaction ^b^ |
|  | |  | | FA <600 µg | | | | FA ≥600 µg |  |  |
| ALT (n=230) | |  | | n=184 | | | | n=46 |  |  |
|  | | LMWP | | 9.97 (0.52, 19.6) | | | | -3.61 (-20.9, 15.6) |  | p=0.068 |
|  | | HMWP | | 0.69 (-7.92, 10.1) | | | | -9.96 (-24.8, 7.34) |  | p=0.242 |
|  | | Non-Essential Metals | | 3.69 (-8.12, 17.4) | | | | -8.83 (-27.3, 14.2) |  | p=0.397 |
|  | | Essential Metals | | 8.85 (-5.56, 24.1) | | | | -3.51 (-25.9, 21.9) |  | p=0.471 |
|  | | Pesticides | | 3.39 (-5.45, 13.7) | | | | -1.18 (-18.3, 18.3) |  | p=0.775 |
|  | | Air Pollution | | 7.05 (-1.80, 16.6) | | | | -11.7 (-23.7, 3.30) |  | p=0.023* |
|  | | Overall MDC-Mixtures | | 8.20 (-5.81, 25.8) | | | | -12.5 (-33.1, 14.9) |  | p=0.158 |
| AST (n=230) | |  | | |  | n=183 | | n=47 |  |  |
|  | | LMWP | | 3.51 (-3.33, 10.4) | | | | -9.13 (-20.8, 4.75) |  | p=0.087 |
|  | | HMWP | | 0.68 (-5.48, 7.22) | | | | -11.0 (-22.2, 1.69) |  | p=0.126 |
|  | | Non-Essential Metals | | 1.40 (-7.01, 9.57) | | | | -5.67 (-21.2, 13.7) |  | p=0.433 |
|  | | Essential Metals | | 6.19 (-3.65, 17.0) | | | | -1.90 (-20.7, 19.6) |  | p=0.530 |
|  | | Pesticides | | 2.79 (-3.15, 9.14) | | | | -9.48 (-21.0, 3.59) |  | p=0.116 |
|  | | Air Pollution | | 3.54 (-2.21, 9.84) | | | | -4.03 (-15.1, 8.50) |  | p=0.428 |
|  | | Overall MDC-Mixtures | | 4.15 (-5.29, 14.6) | | | | -13.8 (-29.8, 6.80) |  | p=0.118 |
| **P*-value<0.05 | | | | | | | | | | |
| ^a^ Effect estimates for the associations between MDC-mixtures and liver enzymes from BWQS regression models by strata of high vs. low folic acid levels using the clinical recommended average folic acid supplementation cut-off of 600 µg for pregnant populations. Estimates for continuous outcomes were back-transformed from the log-scale by exponentiating the value, subtracting 1, and multiplying by a 100 and are expressed as % change in the outcome by quartile increase in the MDC-mixture. Estimates and 95% CIs were exponentiated to show ORs for dichotomized outcomes. Models were adjusted for continuous maternal pre-pregnancy BMI, SES, maternal parity, maternal smoking status and alcohol intake during pregnancy, and age at parturition.  ^b^ *P*-value for the cross-product term between dichotomized folic acid supplementation and continuous BWQS cumulative index from linear regression. BWQS cumulative index was extracted from the BWQS model for MDC groups of interest (2 air pollutants for air pollution, 5 pesticides, 7 essential and 14 non-essential elements or metals, 10 high-molecular-weight phthalates, and 5 low-molecular-weight phthalates). Quartiled indexes were averaged across chemicals within MDC groups to estimate the BWQS cumulative mean index.  Abbreviations: folic acid (FA), Alanine Transaminase (ALT), Aspartate Aminotransferase (AST), high-molecular-weight phthalate (HMWP) metabolites, low-molecular-weight phthalates (LMWP) metabolites, and metabolism-disrupting chemicals (MDCs). | | | | | | | | | | |
| **Table S10. Effect Modification by Puberty at Follow-Up Year ~9 in the Association of MDC Mixtures with Liver Injury in PROGRESS Children.** | | | | | | | | | | |
|  |  | |  | Puberty Status Group ^a^ | | | | |  |  |
| Liver Outcome | MDC Mixture | | | Percent Change (95% CIs) | | | Percent Change (95% CIs) | |  | *P*-for interaction ^b^ |
|  |  | | | Pre-puberty | | | Puberty | |  |  |
| ALT (n=203) |  | |  | n=42 | | | n=161 | |  |  |
|  | LMWP | | | 9.10 (-8.68, 31.8) | | | 5.79 (-3.91, 16.9) | |  | p=0.412 |
|  | HMWP | | | 16.3 (-2.26, 38.4) | | | 8.06 (-1.86, 19.0) | |  | p=0.294 |
|  | Non-Essential Metals | | | 13.9 (-13.5, 48.9) | | | 7.34 (-5.69, 22.5) | |  | p=0.766 |
|  | Essential Metals | | | -4.71 (-30.4, 29.7) | | | 5.46 (-10.8, 23.5) | |  | p=0.264 |
|  | Pesticides | | | 37.2 (13.7, 65.8)* | | | 3.73 (-7.33, 15.9) | |  | p=0.196 |
|  | Air Pollution | | | 18.5 (1.10, 38.8)* | | | 5.77 (-4.46, 16.9) | |  | p=0.132 |
|  | Overall MDC-Mixtures | | | 29.7 (-1.53, 70.7) | | | 11.9 (-3.97, 29.4) | |  | p=0.356 |
| AST (n=204) |  | |  | n=43 | | | n=161 | |  |  |
|  | LMWP | | | 9.02 (0.23, 18.8)* | | | 4.60 (-0.57, 9.90) | |  | p=0.480 |
|  | HMWP | | | 11.7 (2.54, 21.9)* | | | 4.89 (-0.45, 10.5) | |  | p=0.456 |
|  | Non-Essential Metals | | | 5.66 (-7.34, 19.7) | | | 4.28 (-2.91, 11.9) | |  | p=0.857 |
|  | Essential Metals | | | 5.14 (-9.39, 21.7) | | | 1.53 (-7.42, 10.6) | |  | p=0.519 |
|  | Pesticides | | | 12.6 (0.77, 24.6)* | | | 1.23 (-4.37, 7.14) | |  | p=0.103 |
|  | Air Pollution | | | 9.23 (1.15, 17.5)* | | | 2.50 (-2.56, 7.88) | |  | p=0.181 |
|  | Overall MDC-Mixtures | | | 17.9 (2.34, 36.4)* | | | 6.80 (-1.57, 15.7) | |  | p=0.533 |
| **P*-value<0.05  ^a^ Effect estimates for the associations between MDC-mixtures and liver enzymes from BWQS regression models by strata of pubertal vs. pre-pubertal children are shown. Puberty in PROGRESS children was captured by the Tanner scale for follow-up year ~9. Trained study pediatricians performed Tanner staging as follows: they Tanner staged pubic hair for both sexes, genitals for boys using an orchidometer, and breast development for girls, and assigned values ranging from 1 to 5, where stage 1 indicated that an individual did not initiate development while stages above 1 indicated puberty onset or sexual maturity.^25,26^ Estimates were back-transformed from the log-scale by exponentiating the value, subtracting 1, and multiplying by a 100 and are expressed as % change in the outcome by quartile increase in the MDC-mixture. Models were adjusted for child age at follow-up year 8, continuous maternal pre-pregnancy BMI, SES, sex, maternal parity, maternal smoking status and alcohol intake during pregnancy, and age at parturition.  ^b^ *P*-value for the cross-product term between dichotomized puberty and continuous BWQS cumulative index from linear regression. BWQS cumulative index was extracted from the BWQS model for MDC groups of interest (2 air pollutants for air pollution, 5 pesticides, 7 essential and 14 non-essential elements or metals, 10 high-molecular-weight phthalates, and 5 low-molecular-weight phthalates). Quartiled indexes were averaged across chemicals within MDC groups to estimate the BWQS cumulative mean index.  Abbreviations: Alanine Transaminase (ALT), Aspartate Aminotransferase (AST), high-molecular-weight phthalate (HMWP) metabolites, low-molecular-weight phthalates (LMWP) metabolites, and metabolism-disrupting chemicals (MDCs). | | | | | | | | | | |

| **Table S11. Effect Modification by Sex in the Association Between MDC Mixtures with Liver Injury in PROGRESS Children.** \| \| \| \| \| \| \|; \| \| \| Sex ^a^ \| \| \| \|; Liver Outcome \| MDC Mixture \| \| \| Percent Change (95% CIs) \| Percent Change (95% CIs) \| \| *P*-for interaction ^b^; \| \| \| \| Female \| Male \| \|; ALT (n=203) \| \| \| n=98 \| \| n=105 \| \|; \| LMWP \| \| \| 7.10 (-5.92, 22.2) \| 9.71 (-2.72, 23.3) \| \| p=0.885; \| HMWP \| \| \| 7.83 (-4.97, 21.2) \| 13.5 (0.94, 27.7) \| \| p=0.440; \| Non-Essential Metals \| \| \| 11.3 (-5.85, 30.7) \| 5.49 (-9.91, 22.6) \| \| p=0.762; \| Essential Metals \| \| \| 1.27 (-23.8, 32.0) \| -3.02 (-20.7, 17.5) \| \| p=0.799; \| Pesticides \| \| \| 6.88 (-6.23, 21.0) \| 9.60 (-5.46, 25.8) \| \| p=0.357; \| Air Pollution \| \| \| 8.29 (-4.74, 22.7) \| 9.72 (-1.65, 22.6) \| \| p=0.916; \| Overall MDC-Mixtures \| \| \| 14.4 (-5.44, 37.4) \| 18.2 (-2.70, 43.9) \| \| p=0.605; AST (n=204) \| \| \| n=99 \| \| n=105 \| \|; \| LMWP \| \| \| 7.05 (-1.21, 15.7) \| 4.32 (-1.44, 10.4) \| \| p=0.507; \| HMWP \| \| \| 5.50 (-1.61, 13.7) \| 6.19 (0.09, 12.1)* \| \| p=0.926; \| Non-Essential Metals \| \| \| 6.83 (-3.24, 18.0) \| 3.64 (-3.69, 11.8) \| \| p=0.741; \| Essential Metals \| \| \| 1.89 (-12.2, 17.0) \| 1.05 (-7.64, 10.4) \| \| p=0.609; \| Pesticides \| \| \| 4.26 (-3.53, 12.7) \| 3.04 (-3.81, 10.0) \| \| p=0.494; \| Air Pollution \| \| \| 3.57 (-3.46, 11.4) \| 4.12 (-1.20, 9.82) \| \| p=0.344; \| Overall MDC-Mixtures \| \| \| 9.82 (-2.18, 22.7) \| 8.47 (-1.53, 18.7) \| \| p=0.829; **P*-value<0.05 ^a^ Effect estimates for the associations between MDC-mixtures and liver enzymes from BWQS regression models by strata of male vs. female sex are shown. Estimates were back-transformed from the log-scale by exponentiating the value, subtracting 1, and multiplying by a 100 and are expressed as % change in the outcome by quartile increase in the MDC-mixture. Models were adjusted for child age at follow-up year ~9, continuous maternal pre-pregnancy BMI, SES, maternal parity, maternal smoking status and alcohol intake during pregnancy, and age at parturition. ^b^ *P*-value for the cross-product term between dichotomized sex and continuous BWQS cumulative index from linear regression. BWQS cumulative index was extracted from the BWQS model for MDC groups of interest (2 air pollutants for air pollution, 5 pesticides, 7 essential and 14 non-essential elements or metals, 10 high-molecular-weight phthalates, and 5 low-molecular-weight phthalates). Quartiled indexes were averaged across chemicals within MDC groups to estimate the BWQS cumulative mean index. Abbreviations: Alanine Transaminase (ALT), Aspartate Aminotransferase (AST), high-molecular-weight phthalate (HMWP) metabolites, low-molecular-weight phthalate (LMWP) metabolites, and metabolism-disrupting chemicals (MDCs). \| \| \| \| \| \| \| | | | | |
| --- | --- | --- | --- | --- |
| **Table S12. Effect Modification by Overweight/Obesity at Year ~9 in the Association Between MDC Mixtures with Liver Injury in PROGRESS Children.** \| \| \| \| \| \| \| \| \| \| \|; \| \| \| \| \| Overweight/Obesity Status ^a^ \| \| \| \| \| \|; Liver Outcome \| MDC Mixture \| \| \| \| Percent Change (95% CIs) \| \| \| Percent Change (95% CIs) \| \| \| *P*-for interaction ^b^; \| \| \| \| \| Underweight/Normal \| \| \| Overweight/Obese \| \| \|; ALT (n=202) \| \| \| \| \| n=111 \| \| \| n=91 \| \| \|; \| LMWP \| \| \| \| 4.27 (-5.66, 14.9) \| \| \| 1.20 (-12.3, 17.5) \| \| \| p=0.766; \| HMWP \| \| \| \| 5.09 (-4.51, 15.4) \| \| \| 6.37 (-8,22, 22.9) \| \| \| p=0.821; \| Non-Essential Metals \| \| \| \| -2.58 (-14.3, 10.5) \| \| \| 14.7 (-3.49, 36.7) \| \| \| p=0.070; \| Essential Metals \| \| \| \| -7.99 (-21.4, 6.96) \| \| \| 7.09 (-15.7, 33.1) \| \| \| p=0.408; \| Pesticides \| \| \| \| 11.0 (-0.45, 23.6) \| \| \| 1.79 (-11.2, 16.9) \| \| \| p=0.391; \| Air Pollution \| \| \| \| 7.83 (-2.57, 19.1) \| \| \| 0.18 (-13.2, 15.0) \| \| \| p=0.403; \| Overall MDC-Mixtures \| \| \| \| 5.19 (-10.1, 23.4) \| \| \| 11.7 (-8.92, 37.3) \| \| \| p=0.574; AST (n=203) \| \| \| \| \| n=112 \| \| \| n=91 \| \| \|; \| LMWP \| \| \| \| 5.20 (-0.54, 11.2) \| \| \| 2.44 (-5.15, 10.6) \| \| \| p=0.632; \| HMWP \| \| \| \| 6.07 (0.36, 12.2)* \| \| \| 3.21 (-4.14, 11.6) \| \| \| p=0.579; \| Non-Essential Metals \| \| \| \| 4.43 (-3.67, 13.5) \| \| \| 2.02 (-7.62, 11.8) \| \| \| p=0.867; \| Essential Metals \| \| \| \| -1.30 (-9.94, 8.32) \| \| \| 0.20 (-10.8, 11.7) \| \| \| p=0.836; \| Pesticides \| \| \| \| 9.22 (2.69, 16.3)* \| \| \| -3.50 (-10.6, 4.26) \| \| \| p=0.043*; \| Non-Imputed Pesticides ^c^ \| \| \| \| 9.19 (0.02, 18.2)* \| \| \| -5.33 (-17.4, 8.43) \| \| \| p=0.092; \| Air Pollution \| \| \| \| 4.81 (-1.43, 11.2) \| \| \| 1.91 (-5.10, 9.84) \| \| \| p=0.645; \| Overall MDC-Mixtures \| \| \| \| 10.5 (0.24, 22.3)* \| \| \| 2.65 (-7.80, 14.5) \| \| \| p=0.461; **P*-value<0.05 ^a^ Effect estimates for the associations between MDC-mixtures and liver enzymes from BWQS regression models by strata of body mass index (BMI) status are shown. Childhood BMI during the year ~9 examination were based on BMI z-scores (World Health Organization, WHO, ages >5) where children 1 standard deviation (SD) above the WHO Growth Reference median were considered overweight. Estimates were back-transformed from the log-scale by exponentiating the value, subtracting 1, and multiplying by a 100 and are expressed as % change in the outcome by quartile increase in the MDC-mixture. Models were adjusted for child age at follow-up year ~9, continuous maternal pre-pregnancy BMI, SES, sex, maternal parity, maternal smoking status and alcohol intake during pregnancy, and age at parturition. 1 observation deleted due to missing BMI data. ^b^ *P*-value for the cross-product term between dichotomized overweight status (overweight/obesity vs. normal/underweight) and continuous BWQS cumulative index from linear regression. BWQS cumulative index was extracted from the BWQS model for MDC groups of interest (2 air pollutants for air pollution, 5 pesticides, 7 essential and 14 non-essential elements or metals, 10 high-molecular-weight phthalates, and 5 low-molecular-weight phthalates). Quartiled indexes were averaged across chemicals within MDC groups to estimate the BWQS cumulative mean index. ^c^ Sensitivity analysis without imputed organophosphate pesticides metabolite data (complete dataset analysis, n=104). Abbreviations: Alanine Transaminase (ALT), Aspartate Aminotransferase (AST), high-molecular-weight phthalate (HMWP) metabolites, low-molecular-weight phthalates (LMWP) metabolites, and metabolism-disrupting chemicals (MDCs). \| \| \| \| \| \| \| \| \| \| \|; (**Table S13. Effect Modification by Pre-Pregnancy Obesity Status in the Association Between MDC Mixtures with Liver Injury in PROGRESS Mothers.** \| \| \| \| \| \| \| \| \|; \| \| \| Obesity Status \| \| \| \| \| \|; Liver Outcome \| MDC Mixture \| \| BMI <25 (Underweight/Normal) \| BMI ≥25 & <30 (Overweight) \| \| BMI ≥30 (Obese) \| *P*-interaction ^b^ \| \|; ALT (n=230) \| \| \| n=98 \| n=89 \| \| n=43 \| Overweight vs Normal \| Obese vs Normal \|; \| LMWP \| \| 9.92 (-3.53, 23.6) \| 2.36 (-9.99, 15.9) \| \| 7.81 (-15.8, 37.2) \| p=0.294 \| p=0.769 \|; \| HMWP \| \| 2.33 (-9.43, 15.7) \| -4.51 (-16.1, 8.76) \| \| -3.61 (-20.5, 16.2) \| p=0.385 \| p=0.591 \|; \| Non-Essential Metals \| \| 7.21 (-9.12, 27.2) \| -1.59 (-16.7, 16.3) \| \| -6.61 (-28.5, 20.2) \| p=0.478 \| p=0.448 \|; \| Essential Metals \| \| 14.3 (-9.88, 39.7) \| -0.33 (-16.4, 19.4) \| \| -3.38 (-29.0, 32.8) \| p=0.536 \| p=0.387 \|; \| Pesticides \| \| 7.92 (-4.92, 22.1) \| 2.62 (-10.2, 17.1) \| \| 0.58 (-20.1, 29.3) \| p=0.843 \| p=0.179 \|; \| Air Pollution \| \| 5.88 (-4.81, 18.7) \| 1.00 (-10.8, 13.8) \| \| 3.47 (-14.7, 25.3) \| p=0.918 \| p=0.992 \|; \| Overall MDC Mixtures \| \| 12.0 (-6.56, 36.1) \| -1.79 (-18.1, 17.1) \| \| -4.39 (-31.3, 30.0) \| p=0.350 \| p=0.322 \|; AST (n=230) \| \| \| n=99 \| n=88 \| \| n=43 \| Overweight vs Normal \| Obese vs Normal \|; \| LMWP \| \| -2.98 (-10.3, 5.12) \| 4.01 (-4.95, 14.6) \| \| 3.76 (-22.9, 31.7) \| p=0.431 \| p=0.688 \|; \| HMWP \| \| -2.62 (-9.80, 4.65) \| -0.99 (-9.96, 7.80) \| \| 1.17 (-15.3, 20.6) \| p=0.928 \| p=0.696 \|; \| Non-Essential Metals \| \| 6.40 (-4.48, 20.5) \| -2.82 (-14.4, 10.1) \| \| -2.73 (-22.6, 23.0) \| p=0.422 \| p=0.517 \|; \| Essential Metals \| \| 6.46 (-8.17, 20.5) \| 1.70 (-10.7, 15.7) \| \| -1.58 (-27.0, 33.6) \| p=0.601 \| p=0.311 \|; \| Pesticides \| \| -0.42 (-7.75, 7.60) \| 4.41 (-5.01, 14.6) \| \| 8.43 (-11.1, 33.2) \| p=0.504 \| p=0.865 \|; \| Air Pollution \| \| 0.45 (-6.40, 7.42) \| 2.45 (-6.53, 12.3) \| \| 4.83 (-11.7, 25.4) \| p=0.513 \| p=0.286 \|; \| Overall MDC Mixtures \| \| 0.86 (-10.4, 13.5) \| 0.61 (-11.6, 15.8) \| \| 2.28 (-22.9, 34.1) \| p=0.943 \| p=0.949 \|; **P*-value<0.05 \| \| \| \| \| \| \| \| \|; ^a^ Effect estimates for the associations between MDC-mixtures and liver enzymes from BWQS regression models by strata of body mass index (BMI) status are shown. Estimates were back-transformed from the log-scale by exponentiating the value, subtracting 1, and multiplying by a 100 and are expressed as % change in the outcome by quartile increase in the MDC-mixture. Models were adjusted for continuous maternal pre-pregnancy BMI (accounting for residual confounding), SES, maternal parity, maternal smoking status and alcohol intake during pregnancy, and age at parturition. \| \| \| \| \| \| \| \| \|; ^b^ *P*-value for the cross-product term between categorized obesity status (obesity, overweight vs. normal/underweight) and continuous BWQS cumulative index from linear regression. BWQS cumulative index was extracted from the BWQS model for MDC groups of interest (2 air pollutants for air pollution, 5 pesticides, 7 essential and 14 non-essential elements or metals, 10 high-molecular-weight phthalates, and 5 low-molecular-weight phthalates). Quartiled indexes were averaged across chemicals within MDC groups to estimate the BWQS cumulative mean index. \| \| \| \| \| \| \| \| \|; Abbreviations: Alanine Transaminase (ALT), Aspartate Aminotransferase (AST), high-molecular-weight phthalates (HMWP) metabolites, low-molecular-weight phthalates (LMWP) metabolites, and metabolism-disrupting chemicals (MDCs). \| \| \| \| \| \| \| \| \|) \| \| \| \| \| \| \| \| \| \| \|; **Table S14. Sensitivity BWQS Analyses for the Associations Between Pregnancy MDC Mixtures and Liver Injury (ALT≥25) in PROGRESS Children (OR [95% CIs] per Quartile Increases in MDC Mixtures). ^a^** \| \| \| \| \| \| \| \| \| \| \|; ALT Elevation (ALT ≥25) \| \| MDC Mixture \| \| OR (95% CIs) \| \| \| \| \| \| \|; \| \| LMWP \| \| 1.73 (0.99, 3.17) \| \| \| \| \| \| \|; \| \| HMWP \| \| 1.87 (1.07, 3.39)* \| \| \| \| \| \| \|; \| \| Non-Essential Metals \| \| 1.91 (0.90, 3.79) \| \| \| \| \| \| \|; \| \| Essential Metals \| \| 0.99 (0.26, 3.14) \| \| \| \| \| \| \|; \| \| Pesticides \| \| 1.46 (0.78, 2.65) \| \| \| \| \| \| \|; \| \| Air Pollution \| \| 1.79 (1.07, 3.11)* \| \| \| \| \| \| \|; \| \| Overall MDC-Mixtures \| \| 2.65 (1.14, 6.78)* \| \| \| \| \| \| \|; **P*-value<0.05 \| \| \| \| \| \| \| \| \| \| \|; ^a^ Effect estimates for the associations of interest between MDC-mixtures and liver injury defined as an ALT level above 25 U/L from BWQS regression models. Estimates and 95% CIs were exponentiated to show ORs for dichotomized outcomes. All models were adjusted for child age at follow-up year ~9, continuous maternal pre-pregnancy BMI, SES, maternal parity, maternal smoking status and alcohol intake during pregnancy, and age at parturition. Abbreviations: Alanine Transaminase (ALT), high-molecular-weight phthalates (HMWP) metabolites, low-molecular-weight phthalates (LMWP) metabolites, and metabolism-disrupting chemicals (MDCs). \| \| \| \| \| \| \| \| \| \| \| | | | | |
| **Table S15. Sensitivity BWQS Analyses for the Associations Between Pregnancy Pesticide Mixture and Liver Enzymes in PROGRESS Restricted to Participants with Measured Pesticide Data (% Change [95% CIs] per Quartile Increases in Pesticide Mixtures). ^a^** | | | | |
|  | Liver Outcome | | Main Analyses  Percent Change  (95% CIs) ^a^ | Restricted Analyses  Percent Change  (95% CIs) ^a,b^ |
| **Children** |  |  |  |  |
|  | ALT (n=104) | | 8.33 (-1.11, 18.4) | -0.57 (-13.8, 14.7) |
|  | AST (n=104) | | 3.36 (-1.45, 8.26) | 1.63 (-5.47, 9.63) |
| **Mothers** |  | |  |  |
|  | ALT (n=115) | | 2.58 (-5.08, 10.3) | 10.7 (-1.76, 26.4) |
|  | AST (n=115) | | 1.11 (-4.36, 6.80) | 3.42 (-6.02, 13.3) |
| **P*-value<0.05 | | | | |
| ^a^ Effect estimates for the associations of interest between MDC-mixtures and liver enzymes from BWQS regression models. Continuous estimates were back-transformed from the log-scale by exponentiating the value, subtracting 1, and multiplying by a 100 and are expressed as % change in the outcome by quartile increase in the MDC-mixture. Models in children were adjusted for child age at follow-up year ~9, continuous maternal pre-pregnancy BMI, SES, maternal parity, maternal smoking status and alcohol intake during pregnancy, and age at parturition. Models in mothers were adjusted for continuous maternal pre-pregnancy BMI, SES, maternal parity, maternal smoking status and alcohol intake during pregnancy, and age at parturition. ^b^ Additionally restricted to individuals with available pesticide data. Abbreviations: Alanine Transaminase (ALT), Aspartate Aminotransferase (AST). **Table S16. Sensitivity BWQS Analyses for the Associations Between Pregnancy MDC Mixtures and Liver Injury in PROGRESS Children controlling for Lifestyle Factors (% Change or OR [95% CIs] per Quartile Increases in MDC Mixtures). ^a^** \| \| \| \| \| \| \| \|; Liver Outcome \| MDC Mixture \| \| \| \| \| \| \|; \| \| Percent Change or OR (95% CIs) ^a^ \| \| Percent Change or OR (95% CIs) ^b^ \| \| \| \| Percent Change or OR (95% CIs) ^c^; ALT (n=203) \| \| \| \| \| \| \| \|; Continuous \| HMWP \| 10.1 (1.67, 19.4)* \| \| 9.90 (1.53, 18.9)* \| \| \| \| 9.08 (0.48, 19.1)*; \| Air Pollution \| 9.66 (1.05, 19.6)* \| \| 10.0 (1.48, 19.6)* \| \| \| \| 11.9 (2.58, 22.1)*; Dichotomized (90^th^ percentile) \| HMWP \| 1.94 (1.11, 3.56)* \| \| 1.92 (1.08, 3.47)* \| \| \| \| 1.99 (1.04, 3.99)*; AST (n=204) \| \| \| \| \| \| \| \|; Continuous \| LMWP \| 4.98 (0.73, 9.75)* \| \| 5.03 (0.58, 9.57)* \| \| \| \| 5.05 (0.29, 9.97)*; \| HMWP \| 5.27 (0.80, 10.1)* \| \| 5.04 (0.58, 9.82)* \| \| \| \| 4.71 (-0.06, 9.58); **P*-value<0.05 \| \| \| \| \| \| \| \|; ^a^ Effect estimates for the associations of interest between MDC-mixtures and liver enzymes from BWQS regression models. Estimates for continuous outcomes were back-transformed from the log-scale by exponentiating the value, subtracting 1, and multiplying by a 100 and are expressed as % change in the outcome by quartile increase in the MDC-mixture. Estimates and 95% CIs were exponentiated to show ORs for dichotomized outcomes. All models were adjusted for child age at follow-up year ~9, continuous maternal pre-pregnancy BMI, SES, maternal parity, maternal smoking status and alcohol intake during pregnancy, and age at parturition. ^b^ Model additionally adjusted for sugar-sweetened beverages at ~9 years of age. 2 missing observations without sugar-sweetened beverage data. There were n=201 for ALT analyses and n=202 for AST analyses. ^c^ Model additionally adjusted for average sedentary time at 4 and 6 years (in hours). 21 missing observations without sedentary time data. There were n=182 for ALT analyses and n=183 for AST analyses. Abbreviations: Alanine Transaminase (ALT), Aspartate Aminotransferase (AST), high-molecular-weight phthalates (HMWP) metabolites, low-molecular-weight phthalates (LMWP) metabolites, and metabolism-disrupting chemicals (MDCs). \| \| \| \| \| \| \| \| | | | | |
